# Demographic characteristics and prevalence of asymptomatic *Leishmania donovani* infection in migrant workers working in an endemic area in Northwest Ethiopia

**DOI:** 10.1101/2024.01.08.24300972

**Authors:** Mulat Yimer, Yegnasew Takele, Endalew Yizengaw, Endalkachew Nibret, Petra Sumova, Petr Volf, Gizachew Yismaw, Michael Alehegn, Aileen Rowan, Ingrid Müller, James A. Cotton, Lloyd A. C. Chapman, Pascale Kropf

## Abstract

Visceral leishmaniasis (VL), a neglected tropical disease that causes substantial morbidity and mortality, is a serious health problem in Ethiopia. Infections are caused by *Leishmania* (*L*.) *donovani* parasites. Most individuals remain asymptomatic, but some develop VL, which is fatal if not treated.

We identified the area of Metema-Humera in Northwest Ethiopia as a setting in which we could follow migrant workers when they arrived in an endemic area. The demographic characteristics of this population and factors associated with their risk of asymptomatic infection are poorly characterised. We divided our cohort into individuals who visited this area for the first time (first comers, FC) and those who had already been in this area (repeat comers, RC). We followed them from the beginning (Time 1, T1) to the end of the agricultural season (Time 2, T2), performing tests for sand fly bite exposure (anti-sand fly saliva antibody ELISA) and serology for *Leishmania* infection (rK39 rapid diagnostic test and the direct agglutination test) at each time point and collecting information on risk factors for infection.

Our results show that most migrant workers come from non-endemic areas, are male, young (median age of 20 years) and are farmers or students.

At T1, >80% of them had been already exposed to sand fly bites, as shown by the presence of anti-saliva antibodies. However, due to seasonality of sand flies there was no difference in exposure between FC and RC, or between T1 and T2. The serology data showed that at T1, but not at T2, a significantly higher proportion of RC were asymptomatic. Furthermore, 28.6% of FC became asymptomatic between T1 and T2. Over the duration of this study, one FC and one RC developed VL.

In multivariable logistic regression of asymptomatic infection at T1, only age and the number of visits to Metema/Humera were significantly associated with asymptomatic infection.

A better understanding of the dynamics of parasite transmission and the risk factors associated with the development of asymptomatic infections and potentially VL will be essential for the development of new strategies to prevent leishmaniasis.

## INTRODUCTION

Visceral leishmaniasis (VL) is one of the most neglected of tropical diseases. It is endemic in over 60 countries, with 7 countries (Brazil, Ethiopia, India, Kenya, Somalia, Sudan and South Sudan) accounting for over 70% of the global VL burden (1). An estimated 12,773 new cases of VL were reported in 2022 (2). However, the real number of VL cases is likely to be significantly higher, as there is still a lack of appropriate surveillance in most VL-endemic countries. VL is a major health problem worldwide, resulting in significant economic burden and an estimated annual loss of 4.2 million disability-adjusted life years in 2013 (3). Whereas VL incidence has decreased considerably in India, it is currently relatively stable in Africa (2). In Ethiopia, where this study took place, VL is one of the most significant vector-borne diseases, with over 3.2 million people at risk of infection (4). VL is a growing health problem, with spreading endemic areas and persistently high incidence in existing endemic areas since 2009 (5).

In Ethiopia, the *Leishmania* (*L.*) *donovani* species complex is the causative agent of visceral leishmaniasis. Most individuals infected with *L. donovani* will control parasite replication and will not develop VL. However, some individuals will develop VL, a progressive disease that is characterised by uncontrolled parasite replication in spleen, liver and bone marrow, high levels of inflammation and severe immune dysfunction (1). It is generally fatal if untreated. The main treatment in Ethiopia is a combination of sodium stibogluconate (SSG, 20mg/kg/day) and paromomycin (PM, 15 mg/kg/day) for 17 days and has shown an efficacy of 91.4%(6).

The largest VL-endemic area in Ethiopia is in the northwest, in the Metema/Humera region. However, before 1970, only a few cases of VL were reported in this area (7). A study performed in the 70s showed that 45.6% of farmers were leishmanin skin test positive vs 8.3% positive in the non-farming community (8). Studies published in 1978 and 1979 reported VL patients, describing their clinical features in detail and showed that this disease was associated with a high risk of mortality (9, 10). The study by Mengesha and Abuhoy (9) reported that whereas VL had been known to be endemic in the area of Sudan bordering Northwest Ethiopia, little was known about VL in the Metema/Humera area; despite the fact that the number of VL cases was already increasing in the nearby hospital of Gondar. The sharp increase in VL cases in this area coincided with deforestation (11) and the creation of large, intensively cultivated farms, where crops such cotton, maize and sesame are grown at a commercial scale (12, 13). This fertile area attracts now hundreds of thousands of migrant workers (MW) during the agricultural season, mainly from the neighbouring districts of Amhara and Tigray highlands. Most MW come from the highlands, which are largely non-endemic areas (4). It has been shown that individuals living in VL-endemic areas can develop immunity against *L. donovani* and therefore may be protected against VL (14-17). Due to the lack of exposure to *L. donovani*, MW non-endemic areas are at higher risk of developing VL and indeed, we have recently shown that the large majority of patients treated for VL in Gondar were MW coming from the Metema/Humera area (18). The higher risk of developing VL has been worsened by the HIV pandemic, as HIV increases the risk of developing VL (19, 20). VL/HIV co-infected patients suffer from frequent treatment failure, VL relapses and high morbidity and mortality (18, 21). In Northwest Ethiopia, up to 30% of VL patients are co-infected with HIV (7).

MW arrive in the area from around May, for land clearing, ploughing, sowing and weeding, and may stay until the end of the harvest, in December; some return home in August to attend to their own farm and may come back to the Metema/humera area for the harvest season (12, 13). Agriculture is a significant part of Ethiopia’s economy, accounting for 40% of the gross domestic product (GDP), 80% of exports and an estimated 75% of the country’s workforce (22).

*L. donovani* is transmitted to humans by the bite of infected sand flies. In the Metema/Humera area, *Phlebotomus (P.) orientalis* is the main vector of *L. donovani* (23-26). This species of sand flies is found in habitats where *Balanites aegyptica*, *Acacia seyal* trees, deep cracked soil and termite mounds are present, all are common in this area of Ethiopia (24). These ecological conditions are therefore favourable for the transmission of *L. donovani*.

It has been clearly established that the majority of *L*. *donovani* infections do not lead to VL and remain asymptomatic (27, 28). Different ratios of asymptomatic to symptomatic infection have been shown, for example it was estimated at 5.6:1 in Ethiopia (29) and from 1.6:1 and 2.4:1 in Sudan (30). “Asymptomatic infection” is still not clearly defined (28, 31, 32), but is usually characterised by one or a combination of the following positive tests: serological tests, polymerase chain reaction (PCR) and cellular tests; in individuals who do not show any signs of disease and remain healthy (28, 32). However, at least one study has characterised individuals who had a past history of VL as asymptomatic (33). Two recent reviews (28, 32) have described in detail the different combinations of tests used to attempt to identify cohorts of asymptomatic individuals in endemic areas and both highlight the inadequacy of the currently used tests.

The most commonly used serological tests are the Direct Agglutination Test (DAT), a semi-quantitative test that detects the levels of anti-*Leishmania donovani* antibodies; and a rapid diagnostic test (RDT) rK39 detecting antibodies against a highly conserved region from *L. infantum/chagasi,* rK39, as well as an rK39 ELISA. However, these serological tests have been designed for the diagnosis of VL, and not asymptomatic infection, and are therefore unlikely to be sensitive enough to detect infection in asymptomatic individuals, unless they have high antibody titres. Other tests that measure the adaptive immune response to *Leishmania* parasites are also used, such as the leishmanin skin test (LST), where *Leishmania* antigen is injected intradermally, and a Delayed-type hypersensitivity (DTH) reaction is used as a readout system (34); as well as activation of peripheral blood mononuclear cells (35) or whole blood cells (36) with *Leishmania* antigen. Both PCR and qPCR are also used (28, 32).

In Ethiopia, few studies have estimated the incidence of asymptomatic infection, and all have used different tests or combinations of tests to define “asymptomatic individuals”. In Northwest Ethiopia, van Griensven *et al.* (37) showed a prevalence of *Leishmania* infection of 7.2% in HIV+ individuals, as measured by a positive result for at least one of the following tests: rK39, DAT, PCR or KATex, a latex agglutination test that detects *Leishmania* antigens in urine.

In Benishangul-Gumuz, Western Ethiopia, a cross-sectional study (38) identified different rates of asymptomatic individuals depending on the tests used: 3.2% were positive by rK39, 6.0% by LST, and 5.9% by DAT. In another study performed in the Raya Azebo District of Northeastern Ethiopia (39), 9.1% of males were positive by LST. In the Libo Kemkem and Fogera districts, Northwestern Ethiopia (40), a study showed that 1.5% of individuals were positive by rK39, 5.3% DAT and 5.6% by LST. In Northwest Ethiopia (40), a study in children showed that 9.9% of children were positive by at least one of the following tests: rK39, DAT or LST. Two studies evaluated the incidence of asymptomatic infection in MW working in the Metema/Humera area. The most detailed study to date of seroprevalence in this population is that of Lemma *et al*. (12): out of 359 MW, 12.5% were positive by DAT. Another cross-sectional study tested 185 individuals by rK39 and show that 7.6% were positive (41).

In the current study, we recruited a cohort of MW working in farms in the Metema/Humera area that we divided into MW coming to this area for the first time and MW who had already worked in this area in previous years. We recruited them at the beginning of the agricultural season, to assess how many were already exposed to *L. donovani*; and followed them to the end of the agricultural season to assess how many became exposed to *L. donovani* and how many developed VL. We also collected detailed demographic information for the cohort.

## MATERIALS AND METHODS

### Ethical approvals

This study was approved by the Research and Ethical Review Committee of the College of Science, Bahir Dar University (PGRCsVD/053/2011), the National Research Ethics Review Committee (ref. No 04/2.46/62/61) and Imperial College Research Ethics Committee (ICREC 19IC5110). Informed written consent was obtained from each participant.

### Study area

This prospective study took place in the district of West Armachiho, Northwest Ethiopia (13°59’N latitude and 38° 27’ longitude), at the border with Sudan. The area of this district is 2,465.1km^2^, with altitudes varying from 620 to 850m above sea level. The mean minimum and maximum temperatures during the rainy season vary from 20-35°C and can get as high as 45°C during the dry season. The mean annual rainfall ranges from 850 to 1100mm. The natural vegetation of this district is predominantly *Balanites aegyptica* and *Acacia seyal* trees and the soil is mainly clay and of sandy loam texture. Corn, sorghum, cotton and sesame are the most important cash crops grown in this area (42). There are many private owned farms in the area, offering numerous job opportunities for migrant workers.

The study area was 410 km away from Bahir Dar city. The health post in Korhumer, one of the kebeles in the West Armachiho district, was used to collect blood samples from study participants. Study participants were recruited from nine farms, each farm was on average 10 to 20 km away from the health post in Korhumer.

### Migrant workers recruitment

For the first time point at the beginning of the agricultural season (T1), visits to the nine farms were organised with the manager of each farm. The main aim of this study was to focus on MW who were visiting this endemic area for the first time (first comers, FC). We also recruited individuals who had previously visited this area (repeat comers, RC). Exclusion criteria were individuals <18 years old and individuals with a previous history of VL. Phone numbers, as well as permanent places of residence were collected in the questionnaire. For the second time point at the end of the agricultural season (T2), MW were contacted by phone or via the district health offices or health extension workers, who are health professionals visiting every residence in their catchment area. They came to a health facility near to where they lived or worked to have their blood collected.

### Sample collection

Following a finger prick with a sterile lancet, a drop of blood was collected with a plastic pastette and immediately used to detect the presence of anti*-Leishmania* antibodies by rK39. A further 5 drops of blood were collected and were placed on Whatman No3 filter paper and left to dry. Once dried, they were packed individually in sealable plastic bags, to be used at a later time point for the DAT test. Two ml of venous blood were drawn in heparin tubes, 1 ml was frozen immediately to be used later on for the detection of *Leishmania* DNA by qPCR; and one ml was centrifuged, the plasma was collected and immediately frozen to be used at a later time point to measure the levels of antibodies against *Phlebotomus orientalis* salivary proteins (43).

### Body mass index and middle upper arm circumference

Body mass index (BMI) was measured by dividing bodyweight (kg) by the square of height (m). The middle arm circumference was measured in cm using a non-stretchable tape on a straight arm in the middle between the top of the shoulder and the tip of the elbow.

### Serological tests

The rK39 rapid immunochromatographic test for the detection of antibodies against *Leishmania* species (IT LEISH #710124, Biorad) was used following the manufacturer’s instructions. DAT kits were purchased from the Academic Medical Centre at the University of Amsterdam, The Netherlands. DAT is a direct agglutination test: reconstituted *L. donovani* S1 promastigotes antigen reacts directly with anti-*Leishmania* antibodies in blood to form a clear agglutination visible by the naked eye. Five mm circles of dried blood punched from the paper filter were added to 125μL of saline and the protocol was followed as described in the manufacturer’s instructions (44, 45). DAT results were grouped as negative (<1/1600) and positive (≥1/1600).

### Anti-sand fly saliva antibody ELISA

The ELISA with a combination of two *P. orientalis* recombinant salivary proteins mYEL1 and mAG5 as antigens was performed as described in (43) using the plasma of the migrant workers, as well as the plasma of 24 healthy non-endemic controls (HNEC) recruited in the UK. The results are presented as optical density (O.D.). The cut-off value was calculated by using the average O.D. of non-endemic healthy controls + 3 standard deviations.

### Detection of kinetoplast DNA by qPCR

**Detection of kinetoplast DNA by qPCR**

DNA was extracted from 400µL of whole blood by using a fully automated DNA Extraction CyBio FeliX and smart Blood DNA Midi Direct prep (a96) – FX, reagent kit (Analytik Jena GmbH, Germany) following the manufacturer’s protocol and DNA was eluted in 100ul of PCR grade water.

A real-time quantitative PCR (qPCR) was used for detection of *Leishmania donovani* kDNA in blood samples using:

Forward primer (kDNA-CMF, CTTTTCTGGTCCTCCGGGTAGG), reverse primer (kDNA-CMR, CCACCCGGCCCTATTTTACACCAA) and probe (kDNA-CMP, FAM-TTTTCGCAGAACGCCCCTACCCGC-BHQ1 ZEN) (USA) as described in (https://doi.org/10.1128/jcm.42.11.5249-5255.2004).

A primer/probe mix was prepared by mixing primers and probe to achieve a final concentration of 0.6µM kDNA-CMF, 0.6µM KDNA-CMR and 0.4µM kDNA-CMRP. The final master-mix was prepared by mixing 5 µL template DNA with 1µL of primer/probe mix, 0.5µL of BSA (Promega R396A (AcBSA)), 12.5µL of HotStarTaq mix and 6µL of PCR grade water with a final volume of 25µL/reaction.

PCR was performed using BIO-RAD CFX96 real-time PCR (USA) as described in (46) using the following amplification programme: 95°C for 15 min, followed by 45 cycles of 95°C for 5 seconds, 58°C for 20 seconds, 72°C for 30 seconds. Data were analysed using Bio-Rad CFX Maestro 2.3.

### Statistical analysis

Data were evaluated for statistical differences as specified in the legend of each table and figure. The following tests were used: Mann-Whitney, Kruskal-Wallis, Wilcoxon matched-pairs, Fisher exact and Spearman’s rank. Differences were considered statistically significant at p<0.05. ∗=p<0.05, ∗∗=p<0.01, ∗∗∗=p<0.001 and ∗∗∗∗=p<0.0001. Unless otherwise stated, summary statistics given are medians followed by IQR in square brackets. Univariable and multivariable logistic regressions were performed to assess association of asymptomatic infection at the first time point (T1), and seroconversion between the first and second time points (for those who were negative by both rK39 RDT and DAT at T1 and tested on both rK39 RDT and DAT at T2), with various risk factors. Variables were selected in the multivariable models using backwards elimination from a model with all first order terms included, by excluding variables that were not significant at the 5% level in reverse p-value order. Pairwise agreement between different tests at T1 and at T2, for all MW and for FC and RC separately, was assessed using Cohen’s kappa (47) for agreement between two tests, while agreement between all three tests was assessed with Fleiss’ kappa for agreement between multiple tests (48). Kappa values were interpreted using the Landis and Koch scale (49).

## RESULTS

### Characteristics of the migrant workers’ population

The agricultural season in the area of Metema/Humera attracts hundreds of thousands of migrant workers (MW), who arrive from May and may stay until the end of the harvest, in December (Figure 1A).

**Figure 1:**
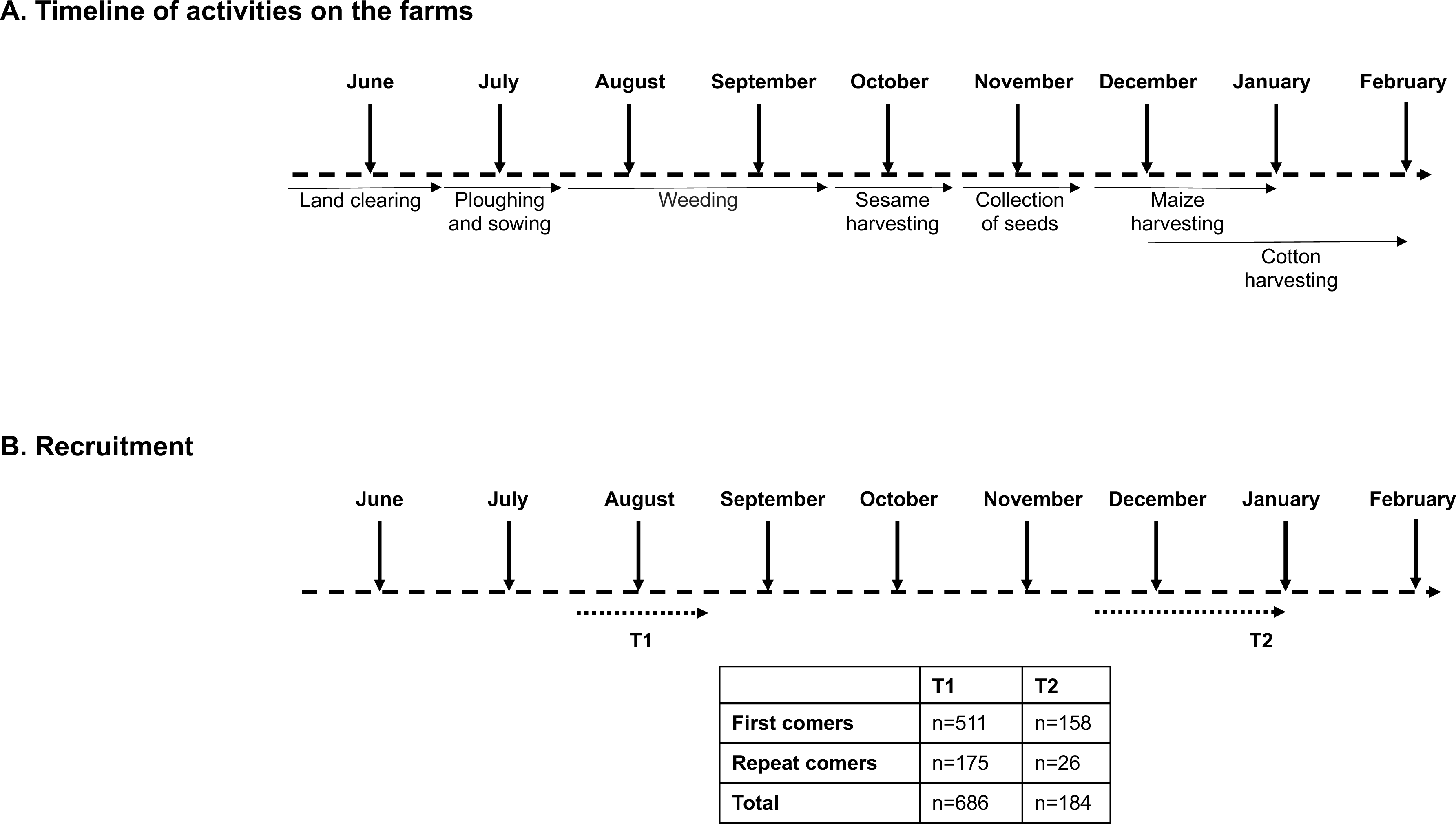
Recruitment and follow-up of migrant workers. (A) Different activities of migrant workers during the agricultural season. (B) 511 migrant workers visiting the area of Metema/Humera for the first time (first comers (FC)) and 175 migrant workers who had already been in this area (repeat comers (RC)) were recruited from mid-July to mid-August. 158 FC and 26 RC were followed-up to the end of the agricultural season from mid-December to the end of January.

In this study, we recruited 686 MW from different farms surrounding Korhumer (Figure 2), at the beginning of the agricultural season (T1, July – August 2019, Figure 1B). Our focus was to assess how many individuals coming for the first time, thus presumably seronegative, seroconverted over time. We therefore recruited a larger number of MW coming for the first time to the endemic area (FC, n=511); and a smaller number of MW who had already visited this area (RC, n=175) (Table 1). 158 FC and 26 RC were followed after 7 months, at the end of the agricultural season (T2, November 2019-January 2020) (Figure 1B).

**Figure 2:**
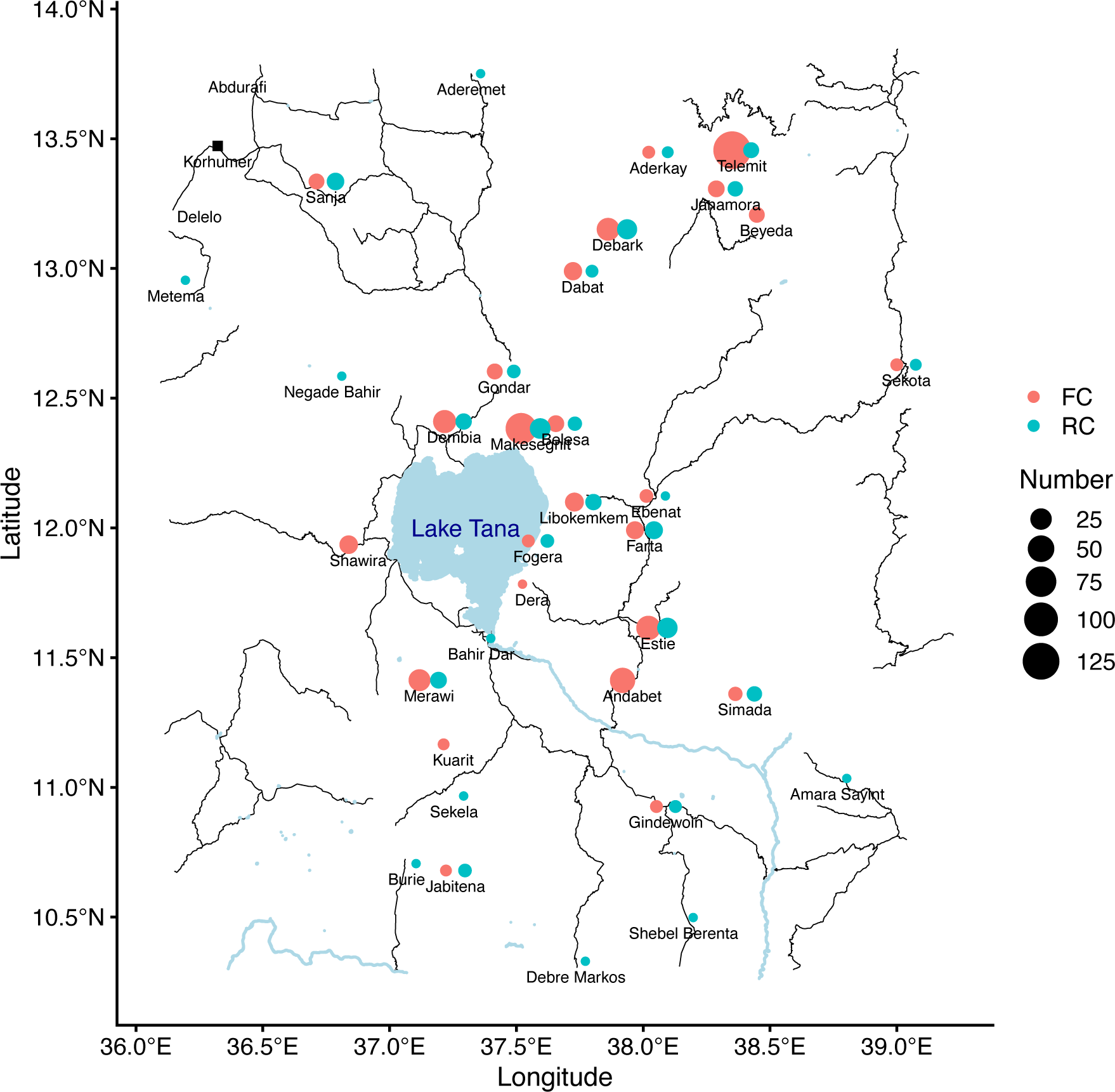
Map of the permanent places of residence of the recruited migrant workers.

**Table 1.**
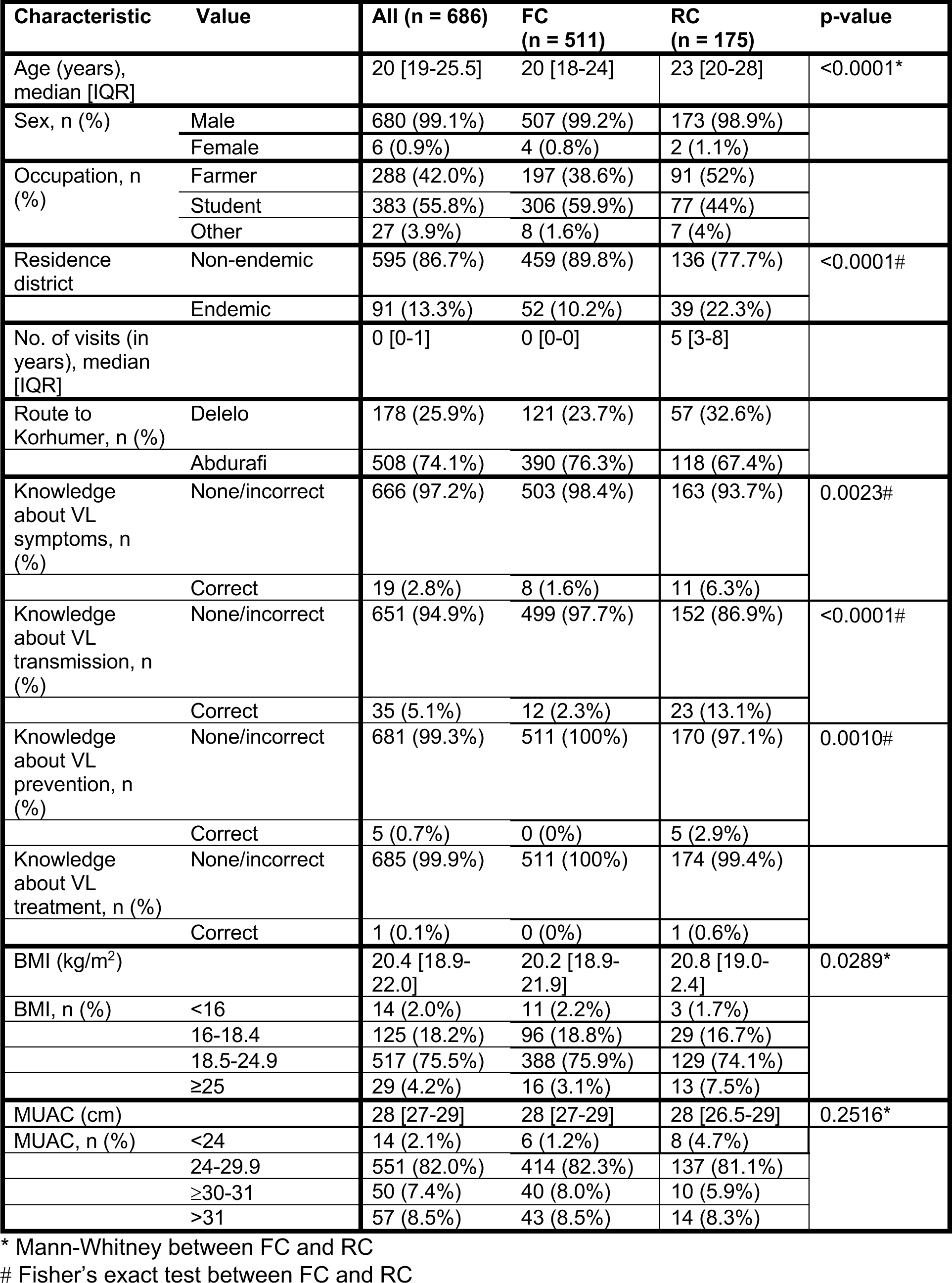
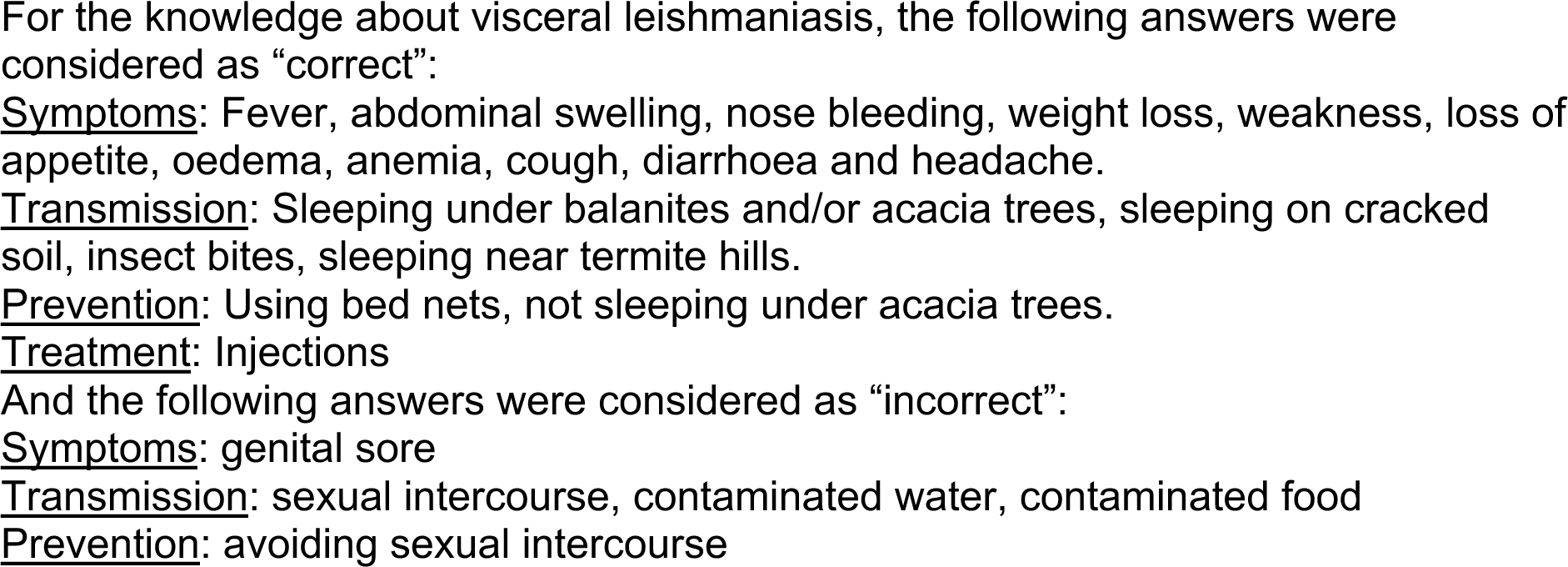
Characteristics of the study population as a whole and split by first comers (FC) and repeat comers (RC).

The median age of the two cohorts was 20 [19-25.5] (Table 1). The median age of RC was significantly higher than FC (23 [20-28] and 20 [18-24], p<0.0001, Table 1, Figure S1A). The majority of FC and RC belong to the 18-29 years age group (Figures S1B and S1C). The large majority were male (n=680, 99.1%, 507 FC and 173 RC). Overall, most FC and RC were farmers and students (Table 1, Figure S2A). The majority of FC were students (59.9%), followed by farmers (38.6%). In contrast, most of the RC were farmers (52%, Table 1, Figure S2A) followed by students (44%, Table 1). The median age of student FC and RC were significantly lower than farmers (Table S1). Most MW (86.7%) came from non-endemic areas (Amhara Regional Health Bureau) (Table 1, Table S2 highlighted in bold, Figure 2). Significantly more RC (n=39, 22.3%) came from areas known to be endemic for VL than FC (n=52 FC, 10.2%, p<0.0001, Table 1). The numbers of times RC visited the VL-endemic area of Metema/Humera to work on farms varied from 1 to 46 times (5 [3-8 times], Table 1, Figure 3). There was a strong positive correlation between the number of visits to the endemic areas and the age of RC (p<0.0001, Figure S1D). Amongst the RC, farmers visited this area significantly more times than students (5.5 [4-8] and 3 [2-7] times, respectively, p<0.0001, Figure S2B). MW recruited in this study came to the farms surrounding Korhumer (Figure 2) via two different routes: Delelo and Abdurafi, the latter is a shorter journey as MW can take transportation directly to Korhumer. In contrast, there is no direct transport to Korhumer from Delelo; this journey has to be done in several stages, and MW have to stop and work on different farms during this journey. The majority of FC and RC travelled via Abdurafi (Table 1).

**Figure 3:**
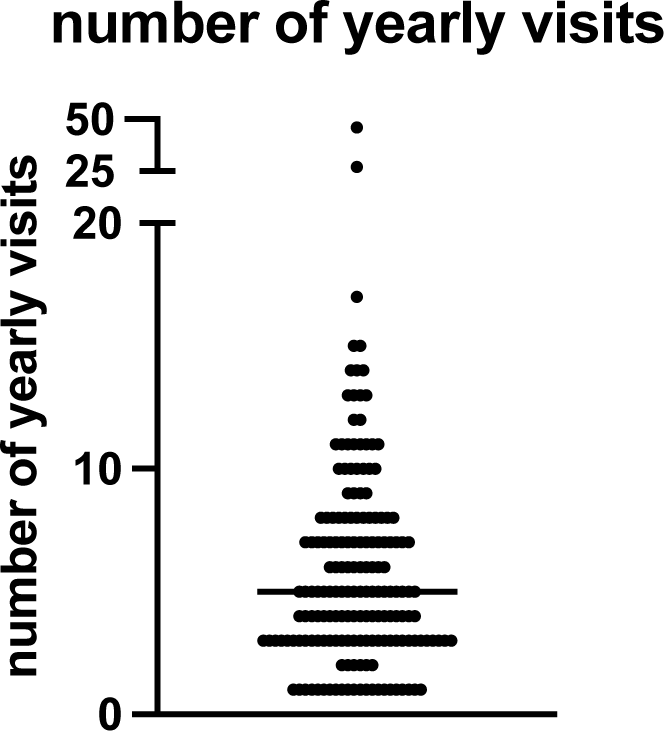
Number of yearly visits to the endemic areas. Number of times (in years) that the RC (n=174) came back to the area of Metema/Humera. The straight line represents the median. RC=repeat comer.

To better evaluate the knowledge of MW about VL, four questions were asked: did they know: i. how the disease is transmitted; ii. how it can be prevented; iii. what the symptoms are; and iv. if the disease can be treated. MW were asked to answer “yes” or “no”, and when they answered “yes”, they were asked to specify (Table 5). A response was considered “correct” when they gave one or more correct answers for each question, as per the Ethiopian National Guidelines (50). As shown in Table 1, the knowledge of MW about VL was poor. This lack of knowledge was even more pronounced for FC: the percentages of FC who knew about transmission, prevention and symptoms were significantly lower than RC (p<0.0001, p=0.0010 and p=0.0023, respectively). Only one MW knew that VL can be treated by injections.

MW were also asked about their exposure to VL risk factors. Out of the 173 RC who were asked at T1 if they slept outside, 170 (98.3%) said yes. 170 (98.3%) said that they did not use bed nets and 3 (1.7%) said they used it “sometimes”. None used repellent. When asked if there were *Balanites aegyptica* and/or *Acacia seyal* trees where they lived or slept, 171 (98.8%) RC said that both were present and 2 (1.2%) had only observed balanites.

Since malnutrition is commonly associated with increased infectious disease susceptibility and severity, we measured the Body Mass Index (BMI) of the two groups at T1. The median BMI of FC was lower than RC (20.2 [18.9-21.9] vs 20.8 [19.0-2.4], p=0.0289), Table 1, Figure S3A). The FC and RC groups were further subdivided into those who were severely malnourished (BMI <16), malnourished (BMI 16-18.4), with a normal BMI (18.5-24.9) or overweight (BMI >25). As shown in Table 1, while the majority of the FC and RC had a normal BMI, 2.2% and 18.8% of FC and 1.7% and 16.7% of RC were severely malnourished or malnourished, respectively. 3.1% of FC and 7.5% of RC were overweight. The median BMI of students was significantly lower as compared to farmers in FC (p<0.0001) and RC (p=0.0117, Figures S3B and S3C). We also measured the medium upper arm circumference (MUAC, Table 1). There was a strong correlation between BMI and MUAC (p<0.0001, Figure S4). As for the BMI measurements, the majority of FC and RC had a normal MUAC (24-30cm, 82% and 82.3%), 1.2% FC and 4.7% RC had a MUAC below 24cm, indicating that they were malnourished and 15.9% FC and 16.5% RC had MUAC measurements >30cm, suggesting that they were overweight.

### Exposure to sand fly saliva

*Phlebotomus orientalis* is the main vector of *L. donovani* and is present in the Metema/Humera area (51). To assess whether the MW had been exposed to sand fly bites, we tested their plasma for the presence of anti-*P. orientalis* saliva antibodies by ELISA (43) at T1 and T2. The O.D. obtained for the MW were significantly higher (Figure 4A, Table 2, p<0.0001) as compared to healthy non-endemic controls (HNEC, 0.0610 [0.0501-0.0766]). Results presented in Figure 4 show that at T1, the majority of FC (85.2%) and RC (81.6%) had antibodies to *P. orientalis* saliva. There was no significant difference in anti-*P. orientalis* antibodies between FC and RC (0.2010 [0.1243-0.5490] and 0.2055 [0.1115-0.3770], respectively, p=0.4125, Figure 4A). For T1, MW were recruited over a period of 4 weeks: results presented in Figure 4B show a positive correlation between the number of days since the start of recruitment and the levels of anti-saliva antibodies (p<0.0001). This positive correlation was true for the FC (p<0.0001), but not the RC (p=0.6979, Figures S5A and S5B). This is likely due to the fact that the majority of FC were recruited during the last part of the first visit while RC were mainly recruited during the first 10 days of the first visit; and indeed, results presented in Figures S5C and S5D show that the levels of anti-saliva antibodies significantly increased between the first (day 1-10) and the second visit (days 20-28) for T1 for FC, but not RC. The levels of anti-saliva antibodies from the individuals coming via Abdurafi, which is the shorter journey to travel to Korhumer, were significantly lower (Figure 4C, p=0.0283).

**Figure 4:**
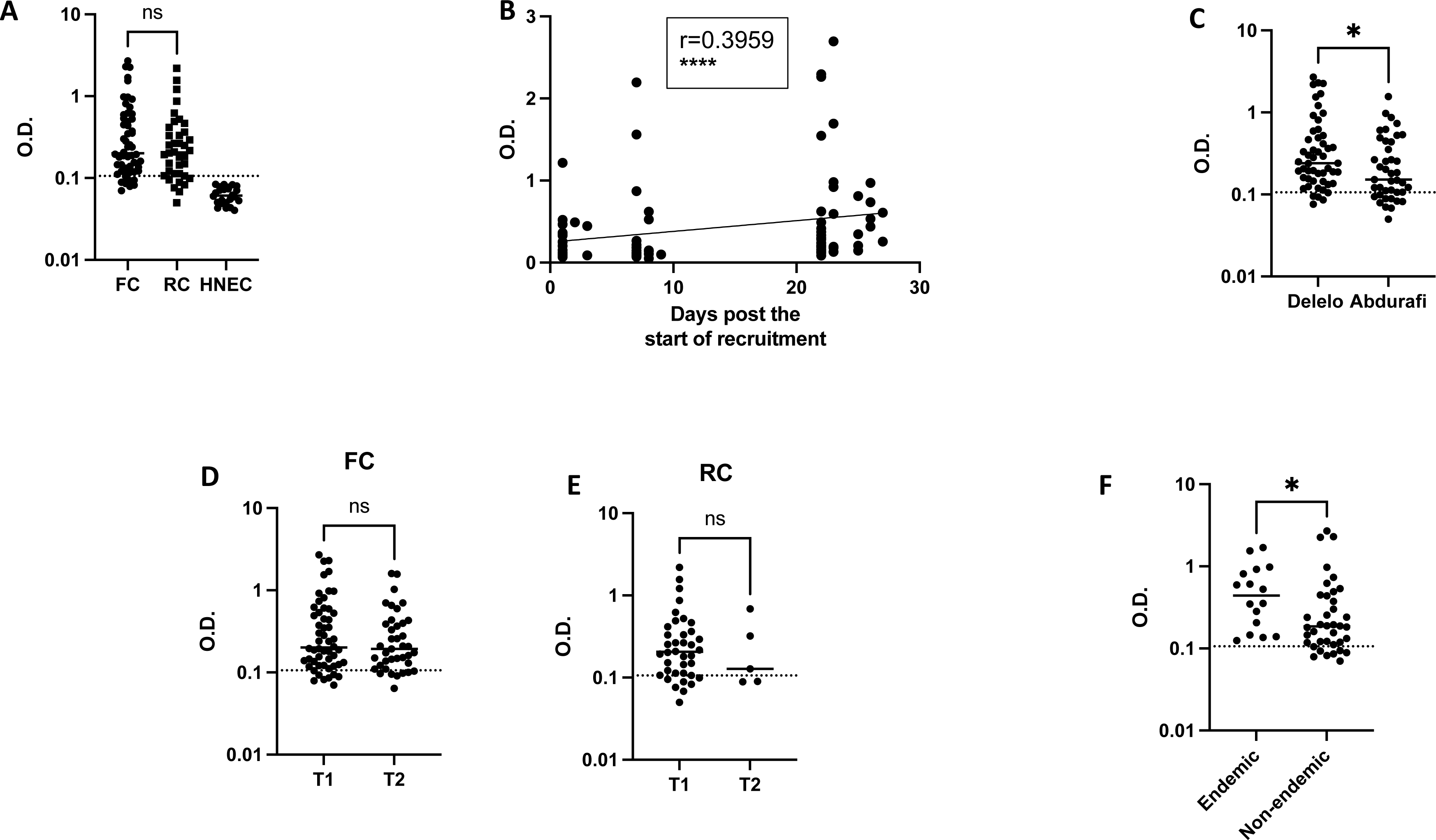
Exposure to sand fly saliva. An ELISA was performed to measure the levels of anti-*P. orientalis* saliva antibodies in plasma as described in (43), the results are presented as optical density (O.D.).

(A) Comparison of FC (n=54), RC (n=38) and Healthy Non-endemic controls (HNEC, n=24). Statistical difference between FC and RC were determined using a Mann-Whitney test. (B) The correlation between the number of days post the start of recruitment and the O.D. was measured by a Spearman test (n=92). (C) Statistical difference between the O.D. of MW who travelled via Delelo (n=51) and those who travelled via Abdurafi (n=41) was determined using a Mann-Whitney test. Statistical differences between T1 and T2 for FC (D) and RC (E) were determined using a Mann-Whitney test. (F) Statistical difference between the O.D. of FC coming from *L. donovani* endemic areas (n=16) and those coming from non-endemic areas (n=38) was determined using a Mann-Whitney test.

The dotted line represents the cut-off value and the straight line represents the median. ns=not significant. FC=first comer, RC=repeat comer.

**Table 2.** Levels of anti-saliva antibodies in the plasma of MW measured by ELISA.

| **T1** | **FC (n = 54)** | **RC (n = 38)** | **p-value\*** | **HNEC (n=24)** | **p-value‡** |
| --- | --- | --- | --- | --- | --- |
| **O.D.** | 0.2010<br>[0.1243-0.5490] | 0.2055<br>[0.1115-0.3770] | 0.4125* | 0.0610<br>[0.0501-0.0766] | <0.0001‡ |
| **T2** | **FC (n = 39)** | **RC (n = 5)** | **p-value** | **HNEC (n=24)** | **p-value** |
| **O.D.** | 0.1930<br>[0.1220-0.3970] | 0.1280<br>[0.0895-0.5035] | 0.3629* | 0.0610<br>[0.0501-0.0766] | <0.0001‡ |
| **p-values^** | 0.5313^ | 0.6224^ |  |  |  |

\* Mann-Whitney between FC and RC
‡ Kruskal-Wallis between FC, RC and HNEC
^ Mann-Whitney between T1 and T2 for FC and RC

There was no significant difference between overall levels of Ab between T1 and T2 for both FC and RC (p=0.5313 and p=0.6224, Figures 4D and E). 26 MW were followed longitudinally, no significant differences in the levels of anti-saliva Ab were observed between T1 and T2 (p=0.5694, Figure S6). At T1, the levels of anti-saliva Ab in the plasma of FC coming from endemic areas (Table 1, in bold) were significantly higher than those coming from non-endemic areas (p=0.0194, Figure 4F). It was not possible to do this comparison with the RC, as no plasma were collected from RC coming from non-endemic areas.

### Serology

Next, to evaluate the number of MW who were seropositive for *Leishmania* infection at T1 and T2 and the number of MW who seroconverted over time, we used two serological tests: rK39 RDT and DAT. We also tested the whole blood for the presence of circulating parasite DNA by using a molecular test.

We first evaluated the numbers of positive FC and RC for each test over time. Results presented in Table 3 show no significant difference in the number of rK39 positive MW between FC and RC at T1, but a significantly higher proportion of RC testing positive by rK39 at T2 as compared to FC.

**Table 3.** Results of different diagnostic tests in first comers (FC) and repeat comers (RC) at time points 1 and 2 (T1 and T2)

|  | Positive<br>n (%) | Negative<br>n (%) | p-value# |
| --- | --- | --- | --- |
| rK39 |  |  |  |
| T1 |  |  |  |
| FC (n=509) | 7 (1.4) | 502 (98.6) | 0.4850 |
| RC (n=175) | 4 (1.8) | 171 (98.2) |  |
| T2 |  |  |  |
| FC (n=158) | 6 (3.8) | 152 (96.2) | 0.0367 |
| RC (n=26) | 4 (15.4) | 22 (84.6) |  |
| DAT |  |  |  |
| T1 |  |  |  |
| FC (n=495) | 10 (2.0) | 485 (98.0) | <0.0001 |
| RC (n=163) | 35 (21.2) | 128 (78.8) |  |
| T2 |  |  |  |
| FC (n=132) | 46 (34.8) | 86 (65.2) | >0.9999 |
| RC (n=22) | 8 (36.4) | 14 (63.6) |  |
| PCR |  |  |  |
| T1 |  |  |  |
| FC (n=47) | 15 (31.9) | 32 (68.1) | 0.3692 |
| RC (n=36) | 15 (41.7) | 21 (58.3) |  |
| T2 |  |  |  |
| FC (n=32) | 16 (50) | 16 (50) | 0.7225 |
| RC (n=10) | 4 (40) | 6 (60) |  |

A significantly higher proportion of RC than FC tested positive by DAT at T1. Figure S7 shows the numbers DAT negative and positive for each titre at T1 and T2.

31.9% FC and 41.7% RC had kinetoplast DNA detected in whole blood by qPCR at T1 and 50% FC and 40% RC at T2. No significant differences between FC and RC were observed by PCR (Table 3).

Next, we assessed the number of FC and RC negative at T1 by rK39, DAT or PCR, who became positive by T2 (Table 4). 151 FC and 22 RC who were negative by rK39 at T1 were followed to T2, none seroconverted. 124 FC and 11 RC who tested negative by DAT at T1 were followed to T2; of those, 39 FC and 2 RC became positive. 14 FC and 7 RC who tested negative by PCR at T1 were followed and at T2, 8 and 2 of these respectively tested positive. The Ct value for each positive sample is shown in Table S3. Of note, at T1, there was no correlation between the number of days post recruitment and the Ct value (p=0.4837, data not shown). Five (38.5%) MW who had detectable kinetoplast DNA at T1 by PCR tested negative by PCR at T2 (Table S4).

**Table 4.**
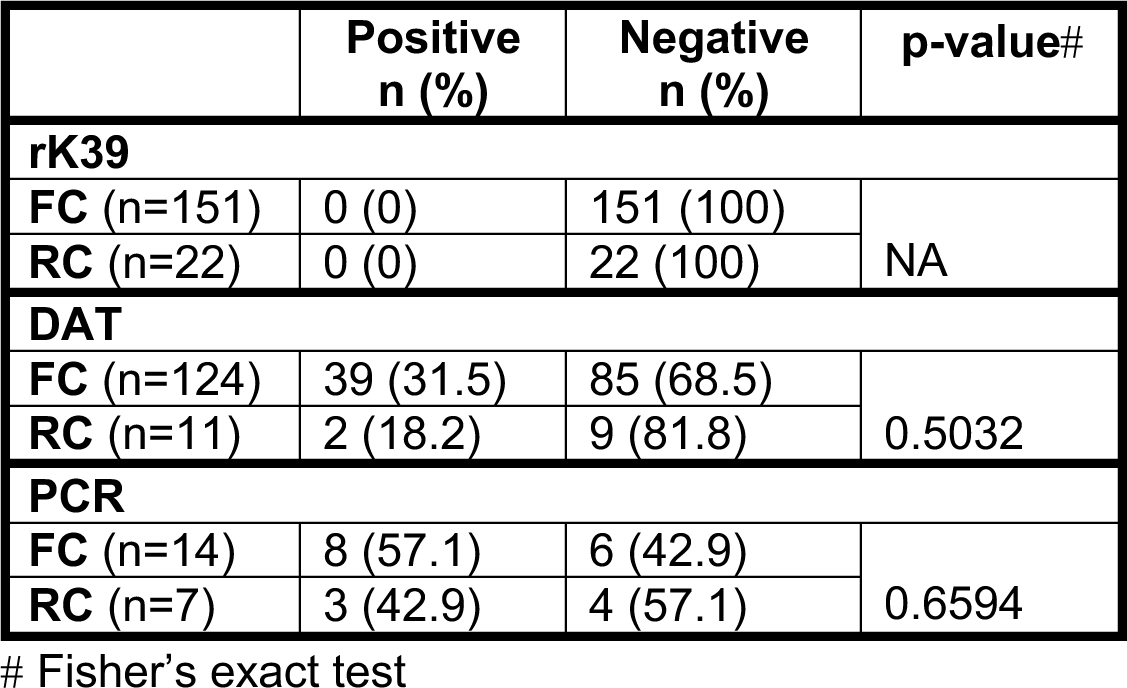
rK39, DAT and PCR conversion between time points 1 and 2 (T1 and T2) for FC and RC.

|  | Positive<br>n (%) | Negative<br>n (%) | p-value# |
| --- | --- | --- | --- |
| rK39 |  |  |  |
| FC (n=151) | 0 (0) | 151 (100) | NA |
| RC (n=22) | 0 (0) | 22 (100) |  |
| DAT |  |  |  |
| FC (n=124) | 39 (31.5) | 85 (68.5) | 0.5032 |
| RC (n=11) | 2 (18.2) | 9 (81.8) |  |
| PCR |  |  |  |
| FC (n=14) | 8 (57.1) | 6 (42.9) | 0.6594 |
| RC (n=7) | 3 (42.9) | 4 (57.1) |  |

### Concordance between positive results

Data on concordance between the different tests at T1 and T2 for all MW, and for FC and RC separately, is presented in Figures S8 and S9 and Cohen’s and Fleiss’ kappa values for agreement between the tests are shown in Tables S5 and S6. In general, the level of agreement between tests was very low, with no FC testing positive on more than one test at T1, and a minority of FC at T2 and RC at T1 and T2 who were tested with two or more tests being positive on both/all tests. The Cohen’s and Fleiss’ kappa values were negative or low (κ < 0.2) for all comparisons for all groups (except for rK39 and DAT results for RC at T2 (κ = 0.56)), indicating no or only slight agreement between tests, albeit with the Cohen’s kappa values being slightly higher for RC than FC at T1, and for rK39 and DAT results vs other pairs of tests at T2.

### Asymptomatic individuals

Due to the lack of clinical evidence for the use of PCR as a diagnostic tool for asymptomatic infections, we identified asymptomatic (AS) FC and RC based on at least one positive test out of the two serological tests performed (rK39 and DAT). We first assessed how many MW were asymptomatic cross-sectionally at T1 and T2 (Table 5). At T1, there were significantly more RC (22.1%) who were AS than FC (3.0%) (p<0.0001). At T2, a higher proportion of FC were AS than at T1: 132 FC were tested with both tests and 34.8% were AS. There was no significant difference in the proportion AS between RC and FC at T2 (Table 5). Out of the 119 FC negative for both rK39 and DAT at T1, 28.6% became asymptomatic by T2. There was no significant difference from RC who became asymptomatic between T1 and T2; however, only 10 were followed to T2 (Table 5).

**Table 5.** Asymptomatic infection at time points 1 and 2 (T1 and T2) and asymptomatic seroconversion between T1 and T2.

|  | Positive<br>n (%) | Negative<br>n (%) | p-value# |
| --- | --- | --- | --- |
| Asymptomatic at T1 |  |  |  |
| FC (n=495) | 15 (3.0) | 480 (97.0) | <0.0001 |
| RC (n=163) | 36 (22.1) | 127 (77.9) |  |
| Asymptomatic at T2 |  |  |  |
| FC (n=132) | 46 (34.8) | 86 (65.2) | >0.9999 |
| RC (n=22) | 8 (36.4) | 14 (63.6) |  |
| Asymptomatic seroconversion from T1 to T2 |  |  |  |
| FC (n=119) | 34 (28.6) | 85 (71.4) | 0.2849 |
| RC (n=10) | 1 (10) | 9 (90) |  |

### MW who developed VL

By T2, 2 individuals developed VL, 1 FC and 1 RC. The trajectory of the different tests between T1 and T2 is shown in Table S7. In addition, one MW called after 3 years to inform us that he had developed VL. He had tested negative by rK39 and DAT at T1 and T2, he was not tested by PCR (data not shown).

### Risk factors

In univariable logistic regression analyses, age and number of visits (in years) were found to be strongly positively associated with odds of asymptomatic infection at the first time point (T1), with each additional year of age giving a 6% (95% CI 3-9%, p<0.0001) increase and each additional visit giving a 26% (95% CI 17-35%, p<0.0001) increase in the odds of asymptomatic infection. Higher BMI was also associated with higher odds of asymptomatic infection, with each unit increase in BMI corresponding to a 16% (95% CI 3-30%, p = 0.014) increase in odds of asymptomatic infection. Being a student was associated with a 56% lower (95% CI 20-75%, p=0.0074) odds of asymptomatic infection at T1 compared with being a farmer. No other variables were statistically significantly associated with odds of asymptomatic infection at T1 (Table 6).

**Table 6.**
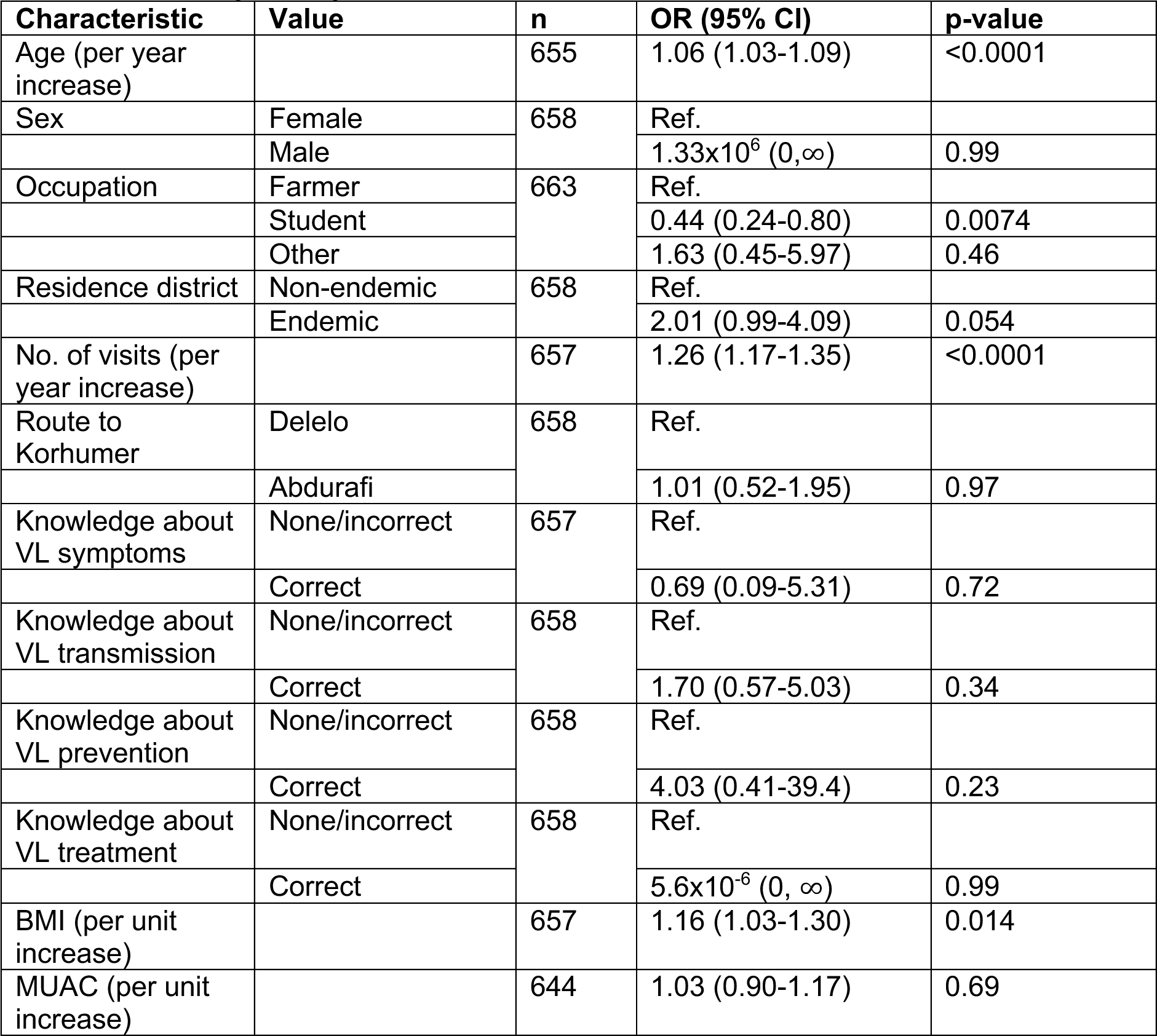
Association of asymptomatic infection at time point 1 (T1) with different risk factors from univariable logistic regressions.

When risk factors were combined in the multivariable logistic regression model, only age and the number of visits to Metema/Humera remained significantly associated with asymptomatic infection at T1, and the effects of each on odds of asymptomatic infection decreased, with each additional year of age corresponding to a 4% (95% CI 0.2-8%) increase in odds, and each additional visit corresponding to a 23% (95% CI 14-32%) increase in odds (Table 7).

**Table 7.** Factors associated with asymptomatic infection at time point 1 (T1) from multivariable logistic regression (n = 639)

| **Characteristic** | **OR (95% CI)** | **p-value** |
| --- | --- | --- |
| Age (per year increase) | 1.04 (1.00-1.08) | 0.039 |
| No. of visits (per year increase) | 1.23 (1.14-1.32) | <0.0001 |

Out of all the risk factors tested in univariable logistic regressions for seroconversion between T1 and T2, only the route to Korhumer was found to be significantly associated with odds of seroconversion, with those coming to Korhumer by the shorter route via Abdurafi having 3.5 (95% CI 1.1-10.7) times higher odds of seroconversion than those coming via Delelo. This remained the only variable significantly associated with seroconversion between T1 and T2 in the multivariable logistic regressions (Table 8).

**Table 8.**
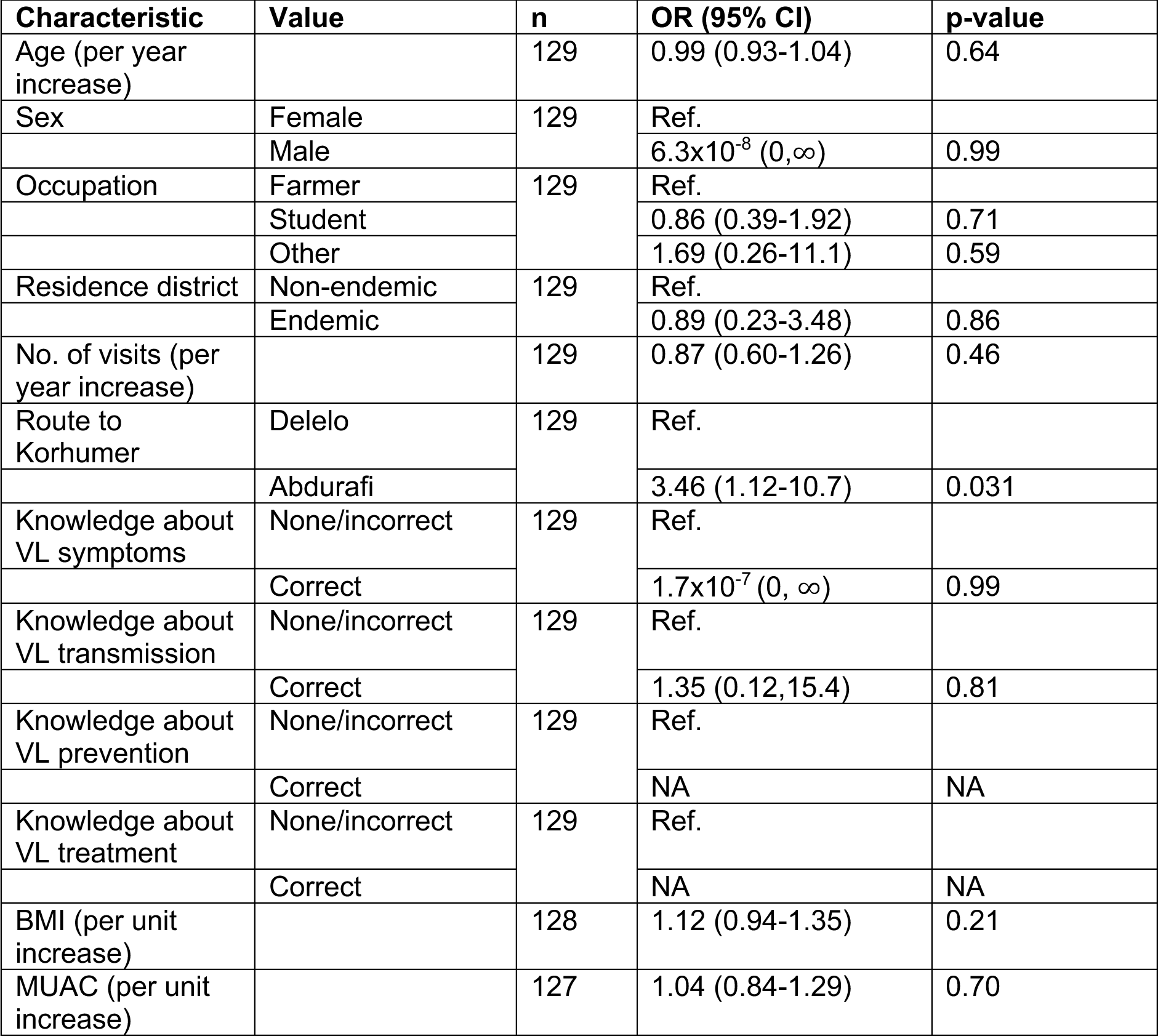
Association of seroconversion between time point 1 (T1) and time point 2 (T2) with different risk factors from univariable logistic regressions.

## DISCUSSION

Our paper is the most detailed socio-demographic and epidemiological study to date of MW working in farms in the West Armachiho district. This is also the first study to compare MW visiting this VL-endemic area for the first time (FC) with MW who have already visited this area (RC) and follow them from the beginning to the end of an agricultural season. To this end, we recruited 511 FC and 175 RC. The majority of FC and RC were aged 18-29 years and most were male as observed in other studies performed in Northwest Ethiopia(12, 52-55)

In our study, we recruited a majority of FC, as our aim was to follow them over time to assess how many might become infected with *L. donovani* and develop VL. Most FC were students whereas the majority of RC were farmers. Other studies have assessed the main occupation of these MW and have also shown that most are farmers (54-56). This is likely due to MW making more money while working on commercial farms than they make on their own farm (13), which may also explain why many of the RC who were farmers had already come several times.

We also assessed knowledge of VL in FC and RC and found that knowledge of MW about transmission, prevention, symptoms and treatment of VL was very poor and considerably worse than reported in previous studies (13, 56). The difference between these studies and our study might be explained by the fact that the majority of our cohort were FC, whose knowledge about this disease was significantly worse than that of RC.

Malnutrition is commonly associated with increased infectious disease susceptibility and severity. It impacts both innate and acquired immunity and can therefore result in immunodeficiencies(57-59). Patients with VL are known to have low BMI (18), but a causal link between malnutrition and risk of developing VL has never been shown. Our results show that >20% of FC and >17% of RC were underweight and that students both in the FC and the RC groups had a significantly lower BMI than the farmers.

To assess the level of exposure to sand fly bites, we measured the levels of antibodies against recombinant salivary proteins from *P. orientalis* (43). Our results show that the majority of FC and RC were positive for antibodies against *P. orientalis* saliva, suggesting that they had been repeatedly exposed to the bites of this sand fly species. There was no difference between the two cohorts at T1. This is likely to be because immunity to sand fly saliva is seasonal and relies on the continued presence of sand flies(60-62), so that RC who go back to non-endemic areas lose their immune response to sand fly salivary proteins. The MW were not asked about the time they had already spent in the farms when they were recruited for this study, but they were likely to have already been there, as our recruitment was started in July, and the agricultural season starts in May/June. Our results collected at T1 show that the longer the MW had stayed in this area, the higher their anti-sand fly saliva antibody levels. However, there was no difference between T1 and T2. This reflects seasonal dynamics of *P. orientalis* which is abundant in May and June but drops in numbers drastically in the wet season (July-September) (63). In addition, this may be a result of levels of these antibodies having reached a plateau, as previously shown (64).

To estimate the prevalence of *L. donovani* infections in the two cohorts of MW, as well as the number of MW who seroconvert over time, we used two serological tests: rK39 RDT and DAT. Both tests were chosen because they are relatively easy to do in the demanding conditions of a field setting. However, as tests designed for the diagnosis of VL, they are probably not sensitive enough to identify all asymptomatic MW, but might be useful to identify those who have high titres. As expected, a small number of FC and RC tested positive by rK39 RDT. No seroconversion of those who tested negative at T1 was observed at T2. Use of a semi-quantitative rK39 ELISA as opposed to the RDT might have identified more asymptomatic individuals (65). Significantly more RC than FC tested positive by DAT at T1. To the best of our knowledge, only one other study has tested MW in the Humera area by DAT and it found that 12.5% had DAT titres ≥1/800 (12). This is almost double the pooled percentage of FC and RC DAT positive in our study (6.8%). However, there were several differences between this study and ours that may explain this difference. In the study by Lemma *et al.,* plasma was used to perform the DAT whereas in our study, we used dried blood spots. We also used a higher threshold for positivity (>1/800) (using the same threshold of ≥1/800, 8.8% of MW would have been deemed DAT positive). And we recruited a much higher proportion of FC than Lemma *et al* due to purposively recruiting FC. A high percentage of FC (31.5%) who tested negative by DAT at T1 seroconverted by T2, indicating that a considerable proportion of FC were infected by *Leishmania* parasites and developed *Leishmania*-specific antibodies. A smaller percentage (18.2%) of RC seroconverted at T2, but caution should be exercised in interpreting this result as only 11 RC negative by DAT at T1 were followed to T2.

We also tested whole blood for the presence of parasite DNA by PCR and our results show that at T1, a high proportion of MW (31.9% FC and 40.7% RC) had kinetoplast DNA detected in whole blood samples by PCR. This was especially unexpected for the FC. Due to the extreme heat during the daytime, MW mainly work during the night, when sand flies are most active. Some MW use frontal lamps and have described that sand flies form “clouds” around their face. It is therefore possible that the high number of PCR-positive individuals is a result of the remarkably high number of sand flies biting MW at night. While there are no data on the percentage of infected sand flies in this endemic area, it is tempting to speculate that this might result in high levels of parasite transmission. Indeed, previous studies showed that levels of anti-saliva antibodies are a marker of *Leishmania* transmission in dogs (66) or humans (67). Quinnell *et al*. also found that this was not predictive of the outcome of the infection (66).

Our results show that a substantial proportion of FC (34.8%) and RC (36.4%) were asymptomatic at T2, and that a high proportion of FC had been asymptomatically infected at T1 (28.6%). There is no gold standard to define asymptomatic infections (28, 32). We based our definition of “asymptomatic infection” on at least one of two serological tests (rK39 and DAT) being positive, but not on PCR positivity. Although there are various PCR assays (68, 69), few have been validated with a large number of clinical samples, especially on asymptomatic cohorts. PCR results can be inconsistent, especially with low parasite load: the study of Abbasi *et al.* showed that over 40% of samples with low parasite load tested negative when retested with the same method (70). Furthermore, no studies have tested longitudinally PCR-positive asymptomatic individuals. And indeed, our results show that almost half of PCR-positive individuals reverted to being PCR negative at T2. Furthermore, as shown by others (71-74), there was poor agreement between DAT and PCR tests. At T2, 16 FC were tested by both PCR and DAT, and only 2 were positive for the presence of parasite DNA in the blood, as well as anti-*Leishmania* antibodies. In addition to the fluctuating nature of PCR results, it is also not possible to conclude from PCR-positive results if *Leishmania* parasites will survive, establish themselves in phagocytic cells and result in an asymptomatic infection or even VL; or if it is DNA from parasites killed by the innate immune response. Only a large-scale longitudinal study with PCR-positive individuals will answer this question.

Although DAT might be a better option to identify asymptomatic individuals, as it measures anti-*Leishmania* antibodies, false positives have been observed (75). Furthermore, due to lot-to-lot variations and possible different interpretations of the results, it is difficult to standardise the use of this test (76).

LST is no longer in use since there was no standardisation of the production of leishmanin. However, a recent paper (77) has described a workflow for the production of GMP-grade leishmanin that could be used for large clinical studies. Other tests such as the whole blood assay (WBA) has been shown to be more specific than the LST (78). However, in our case, it was not possible to use the WBA as we could not maintain the temperature of the incubator at 37°C in ambient temperatures that were >40°C. Extensive longitudinal clinical studies are required to determine if these tests are more appropriate for the identification of asymptomatic individuals.

Out of the 686 MW recruited at the beginning of the agricultural season, 184 were followed until the end of the season and of those, two developed VL. It had been clearly explained to all participants of this study how to get in touch if they developed any symptoms of VL, but except for one individual who called in 2022, no other participants made contact. We cannot ascertain from these data that only three out of the 686 MW developed VL. Any follow-up of this population has been severely hampered by the SARS-CoV-2 pandemic and the political instability in Ethiopia. In a multivariate analysis, age and the number of visits to farms in Metema/Humera were significantly associated with being asymptomatic at T1. This is despite age and number of visits being positively correlated (Figure S1D), and thus suggests that cumulative exposure through age contributes to infection risk in addition to time spent in the VL-endemic area of Metema-Humera. In univariable logistic regressions for seroconversion between T1 and T2, there was some evidence that the route to Korhumer was associated with odds of seroconversion, with those coming to Korhumer by the shorter route via Abdurafi having higher odds of seroconversion. This result should be interpreted with caution given the small numbers involved (only 4 of the 33 individuals who travelled via Delelo became asymptomatic). The results from this study give a clear picture of the demographic and epidemiological profiles of a population of MW, mainly coming from non-endemic areas, who travelled to the endemic area of Metema-Humera during the agricultural season. We show that the majority of these individuals had been repeatedly exposed to sand fly bites. A considerable proportion of individuals who visited these farms for the first time became asymptomatically infected, but few developed VL. Age and the number of visits to this area are major risk factors for asymptomatic infection. A better understanding of the prevalence of asymptomatic infection and incidence of VL, as well as of the transmission dynamics of *Leishmania* parasites, are essential prerequisites for improving the prevention and treatment of visceral leishmaniasis amongst migrant workers in Ethiopia.

## Data Availability

All data produced in the present study are available upon reasonable request to the authors

## Acknowledgements

We are grateful to the staff of the Amhara Public Health Institute for their support and to Friew Mekuanint Wubieneh for his unvaluable help in the recruitment of migrant workers. This research is jointly funded by the UK Medical Research Council (MRC) and the Foreign Commonwealth and Development Office (FCDO) under the MRC/FCDO Concordat agreement (MR/R021600/1) (MY, EY, JAC, LACC, PK). PS and PV were supported by ERD funds, project CeRaViP (16_019/0000759).

## REFERENCES

1. Burza S, Croft SL, Boelaert M. Leishmaniasis. Lancet. 2018;392(10151):951–70.

2. Ruiz-Postigo JA, Jain S, Madjou S, Virrey Agua JF, Maia-Elkhoury AN, Valadas S, et al. Global leishmaniasis surveillance, 2022: assessing trends over the past 10 years. Weekly epidemiological record. 2023;40(98):471–87.

3. DALYs GBD, Collaborators H, Murray CJ, Barber RM, Foreman KJ, Abbasoglu Ozgoren A, et al. Global, regional, and national disability-adjusted life years (DALYs) for 306 diseases and injuries and healthy life expectancy (HALE) for 188 countries, 1990-2013: quantifying the epidemiological transition. Lancet. 2015;386(10009):2145-91.

4. Gadisa E, Tsegaw T, Abera A, Elnaiem DE, den Boer M, Aseffa A, et al. Eco-epidemiology of visceral leishmaniasis in Ethiopia. Parasit Vectors. 2015;8:381.

5. WHO. Number of cases of visceral leishmaniasis reported Data by country 2023 [Available from: https://apps.who.int/gho/data/node.main.NTDLEISHVNUM?lang=en.

6. Musa A, Khalil E, Hailu A, Olobo J, Balasegaram M, Omollo R, et al. Sodium stibogluconate (SSG) & paromomycin combination compared to SSG for visceral leishmaniasis in East Africa: a randomised controlled trial. PLoS Negl Trop Dis. 2012;6(6):e1674.

7. Leta S, Dao TH, Mesele F, Alemayehu G. Visceral leishmaniasis in Ethiopia: an evolving disease. PLoS Negl Trop Dis. 2014;8(9):e3131.

8. Fuller GK, Lemma A, Haile T, Atwood CL. Kala-azar in Ethopia I: Leishmanin skin test in Setit Humera, a kala-azar endemic area in northwestern Ethopia. Ann Trop Med Parasitol. 1976;70(2):147–63.

9. Mengesha B, Abuhoy M. Kala-azar among labour migrants in Metema-Humera region of Ethiopia. Trop Geogr Med. 1978;30(2):199–206.

10. Maru M. Clinical and laboratory features and treatment of visceral leishmaniasis in hospitalized patients in Northwestern Ethiopia. Am J Trop Med Hyg. 1979;28(1):15–8.

11. G. Yeshineh MM, G. Zeleke, G. Desta. Threats and management options of the green belt natural forest, northwest lowlands of Ethiopia. Trees, Forests and People. 2022;9:100305.

12. Lemma W, Tekie H, Yared S, Balkew M, Gebre-Michael T, Warburg A, et al. Sero-prevalence of Leishmania donovani infection in labour migrants and entomological risk factors in extra-domestic habitats of Kafta-Humera lowlands - kala-azar endemic areas in the northwest Ethiopia. BMC Infect Dis. 2015;15:99.

13. Alemayehu M, Paintain L, Adera C, Berhe R, Gebeyehu A, Gizaw Z, et al. Impact of Education on Knowledge and Practice of Kala Azar Preventive Measures among Seasonal and Migrant Agricultural Workers in Northwest Ethiopia. Am J Trop Med Hyg. 2020;102(4):758–67.

14. Ali A, Ashford RW. Visceral leishmaniasis in Ethiopia. I. Cross-sectional leishmanin skin test in an endemic locality. Ann Trop Med Parasitol. 1993;87(2):157–61.

15. Ali N, Mekuria AH, Requena JM, Engwerda C. Immunity to visceral leishmaniasis. J Trop Med. 2012;2012:780809.

16. Rijal S, Sundar S, Mondal D, Das P, Alvar J, Boelaert M. Eliminating visceral leishmaniasis in South Asia: the road ahead. BMJ. 2019;364:k5224.

17. Volpedo G, Pacheco-Fernandez T, Bhattacharya P, Oljuskin T, Dey R, Gannavaram S, et al. Determinants of Innate Immunity in Visceral Leishmaniasis and Their Implication in Vaccine Development. Front Immunol. 2021;12:748325.

18. Takele Y, Mulaw T, Adem E, Shaw CJ, Franssen SU, Womersley R, et al. Immunological factors, but not clinical features, predict visceral leishmaniasis relapse in patients co-infected with HIV. Cell Rep Med. 2022;3(1):100487.

19. Lindoso JA, Cota GF, da Cruz AM, Goto H, Maia-Elkhoury AN, Romero GA, et al. Visceral leishmaniasis and HIV coinfection in Latin America. PLoS Negl Trop Dis. 2014;8(9):e3136.

20. Akuffo H, Costa C, van Griensven J, Burza S, Moreno J, Herrero M. New insights into leishmaniasis in the immunosuppressed. PLoS Negl Trop Dis. 2018;12(5):e0006375.

21. Davidson RN. AIDS and leishmaniasis. Genitourin Med. 1997;73:237–9.

22. USAID. Ethiopia: Agriculture and Food Security. 2023. https://www.usaid.gov/ethiopia/agriculture-and-food-security

23. Moncaz A, Kirstein O, Gebresellassie A, Lemma W, Yared S, Gebre-Michael T, et al. Characterization of breeding sites of Phlebotomus orientalis - the vector of visceral leishmaniasis in northwestern Ethiopia. Acta Trop. 2014;139:5–14.

24. Lemma W, Tekie H, Balkew M, Gebre-Michael T, Warburg A, Hailu A. Population dynamics and habitat preferences of Phlebotomus orientalis in extra-domestic habitats of Kafta Humera lowlands--kala azar endemic areas in Northwest Ethiopia. Parasit Vectors. 2014;7:359.

25. Seblova V, Volfova V, Dvorak V, Pruzinova K, Votypka J, Kassahun A, et al. Phlebotomus orientalis sand flies from two geographically distant Ethiopian localities: biology, genetic analyses and susceptibility to Leishmania donovani. PLoS Negl Trop Dis. 2013;7(4):e2187.

26. Elnaiem DA, Dakein O, Alawad AM, Alsharif B, Khogali A, Jibreel T, et al. Outdoor Residual Insecticide Spraying (ODRS), a New Approach for the Control of the Exophilic Vectors of Human Visceral Leishmaniasis: Phlebotomus orientalis in East Africa. PLoS Negl Trop Dis. 2020;14(10):e0008774.

27. Singh OP, Hasker E, Sacks D, Boelaert M, Sundar S. Asymptomatic Leishmania infection: a new challenge for Leishmania control. Clin Infect Dis. 2014;58(10):1424–9.

28. Ibarra-Meneses AV, Corbeil A, Wagner V, Onwuchekwa C, Fernandez-Prada C. Identification of asymptomatic Leishmania infections: a scoping review. Parasit Vectors. 2022;15(1):5.

29. Ali A, Ashford RW. Visceral leishmaniasis in Ethiopia. IV. Prevalence, incidence and relation of infection to disease in an endemic area. Ann Trop Med Parasitol. 1994;88(3):289–93.

30. Zijlstra EE, el-Hassan AM, Ismael A, Ghalib HW. Endemic kala-azar in eastern Sudan: a longitudinal study on the incidence of clinical and subclinical infection and post-kala-azar dermal leishmaniasis. Am J Trop Med Hyg. 1994;51(6):826-36.

31. Hasker E, Kansal S, Malaviya P, Gidwani K, Picado A, Singh RP, et al. Latent infection with Leishmania donovani in highly endemic villages in Bihar, India. PLoS Negl Trop Dis. 2013;7(2):e2053.

32. Pederiva MMC, Santos SMD, Rivarola LGS, Guerreiro VJ, Lopes KS, Lima Junior M, et al. Asymptomatic Leishmania infection in humans: A systematic review. J Infect Public Health. 2023;16(2):286–94.

33. Mohammed R, Melkamu R, Pareyn M, Abdellati S, Bogale T, Engidaw A, et al. Detection of asymptomatic Leishmania infection in blood donors at two blood banks in Ethiopia. PLoS Negl Trop Dis. 2023;17(3):e0011142.

34. Carstens-Kass J, Paulini K, Lypaczewski P, Matlashewski G. A review of the leishmanin skin test: A neglected test for a neglected disease. PLoS Negl Trop Dis. 2021;15(7):e0009531.

35. de Araujo FF, Lakhal-Naouar I, Koles N, Raiciulescu S, Mody R, Aronson N. Potential Biomarkers for Asymptomatic Visceral Leishmaniasis among Iraq-Deployed U.S. Military Personnel. Pathogens. 2023;12(5).

36. Singh OP, Gidwani K, Kumar R, Nylen S, Jones SL, Boelaert M, et al. Reassessment of immune correlates in human visceral leishmaniasis as defined by cytokine release in whole blood. Clin Vaccine Immunol. 2012;19(6):961–6.

37. van Griensven J, van Henten S, Mengesha B, Kassa M, Adem E, Endris Seid M, et al. Longitudinal evaluation of asymptomatic Leishmania infection in HIV-infected individuals in North-West Ethiopia: A pilot study. PLoS Negl Trop Dis. 2019;13(10):e0007765.

38. Bejano S, Shumie G, Kumar A, Asemahagn E, Damte D, Woldie S, et al. Prevalence of asymptomatic visceral leishmaniasis in human and dog, Benishangul Gumuz regional state, Western Ethiopia. Parasit Vectors. 2021;14(1):39.

39. Tadese D, Hailu A, Bekele F, Belay S. An epidemiological study of visceral leishmaniasis in North East Ethiopia using serological and leishmanin skin tests. PLoS One. 2019;14(12):e0225083.

40. Gadisa E, Custodio E, Canavate C, Sordo L, Abebe Z, Nieto J, et al. Usefulness of the rK39-immunochromatographic test, direct agglutination test, and leishmanin skin test for detecting asymptomatic Leishmania infection in children in a new visceral leishmaniasis focus in Amhara State, Ethiopia. Am J Trop Med Hyg. 2012;86(5):792–8.

41. Ayehu A, Aschale Y, Lemma W, Alebel A, Worku L, Jejaw A, et al. Seroprevalence of Asymptomatic Leishmania donovani among Laborers and Associated Risk Factors in Agricultural Camps of West Armachiho District, Northwest Ethiopia: A Cross-Sectional Study. J Parasitol Res. 2018;2018:5751743.

42. Aznaw A, Fininsa C, Sahile S, Terefe G. Assessment of Sesame Bacterial Blight (Xanthomonas Campestris Pv. Sesami) on Sesame (Sesamum indicum L.) in North Gondar, Ethiopia. ABC Journal of Advanced Research. 2018;7:45–58.

43. Sumova P, Sima M, Spitzova T, Osman ME, Guimaraes-Costa AB, Oliveira F, et al. Human antibody reaction against recombinant salivary proteins of Phlebotomus orientalis in Eastern Africa. PLoS Negl Trop Dis. 2018;12(12):e0006981.

44. Adams ER, Jacquet D, Schoone G, Gidwani K, Boelaert M, Cunningham J. Leishmaniasis direct agglutination test: using pictorials as training materials to reduce inter-reader variability and improve accuracy. PLoS Negl Trop Dis. 2012;6(12):e1946.

45. Schallig HD, Schoone GJ, Kroon CC, Hailu A, Chappuis F, Veeken H. Development and application of ‘simple’ diagnostic tools for visceral leishmaniasis. Med Microbiol Immunol. 2001;190(1-2):69–71.

46. Mary C, Faraut F, Lascombe L, Dumon H. Quantification of Leishmania infantum DNA by a real-time PCR assay with high sensitivity. J Clin Microbiol. 2004;42(11):5249–55.

47. Cohen J. A Coefficient of Agreement for Nominal Scales. Educational and Psychological Measurement. 1960;20(1):37–46.

48. Fleiss JL. Measuring nominal scale agreement among many raters. Psychological Bulletin. 1971;76(5):378–82.

49. Landis JR, Koch GG. The measurement of observer agreement for categorical data. Biometrics. 1977;33(1):159–74.

50. Guidelines for diagnosis, treatment and prevention of leishmaniasis in Ethiopia (2013). https://www.afrikadia.org/wp-content/uploads/2018/08/VL_Guidelines_Ethiopia_2013.pdf

51. Gebresilassie A, Kirstein OD, Yared S, Aklilu E, Moncaz A, Tekie H, et al. Species composition of phlebotomine sand flies and bionomics of Phlebotomus orientalis (Diptera: Psychodidae) in an endemic focus of visceral leishmaniasis in Tahtay Adiyabo district, Northern Ethiopia. Parasit Vectors. 2015;8:248.

52. Gelaye KA, Demissie GD, Ayele TA, Wami SD, Sisay MM, Akalu TY, et al. Low Knowledge and Attitude Towards Visceral Leishmaniasis Among Migrants and Seasonal Farm Workers in Northwest Ethiopia. Res Rep Trop Med. 2020;11:159–68.

53. Argaw D, Mulugeta A, Herrero M, Nombela N, Teklu T, Tefera T, et al. Risk factors for visceral Leishmaniasis among residents and migrants in Kafta-Humera, Ethiopia. PLoS Negl Trop Dis. 2013;7(11):e2543.

54. Berhe R, Spigt M, Schneider F, Paintain L, Adera C, Nigusie A, et al. Understanding the risk perception of visceral leishmaniasis exposure and the acceptability of sandfly protection measures among migrant workers in the lowlands of Northwest Ethiopia: a health belief model perspective. BMC Public Health. 2022;22(1):989.

55. Tilaye T, Tessema B, Alemu K. High asymptomatic malaria among seasonal migrant workers departing to home from malaria endemic areas in northwest Ethiopia. Malar J. 2022;21(1):184.

56. Alemu Gelaye K, Debalke G, Awoke Ayele T, Fekadu Wolde H, Sisay MM, Teshome DF, et al. Occupational Health Problems among Seasonal and Migrant Farmworkers in Ethiopia: A Cross-Sectional Study. Risk Manag Healthc Policy. 2021;14:4447–56.

57. Schaible UE, Kaufmann SH. Malnutrition and infection: complex mechanisms and global impacts. PLoS Med. 2007;4(5):e115.

58. Woodward B. Protein, calories, and immune defenses. Nutr Rev. 1998;56(1 Pt 2):S84-92.

59. Takele Y, Adem E, Getahun M, Tajebe F, Kiflie A, Hailu A, et al. Malnutrition in Healthy Individuals Results in Increased Mixed Cytokine Profiles, Altered Neutrophil Subsets and Function. PLoS One. 2016;11(8):e0157919.

60. Oliveira F, Giorgobiani E, Guimaraes-Costa AB, Abdeladhim M, Oristian J, Tskhvaradze L, et al. Immunity to vector saliva is compromised by short sand fly seasons in endemic regions with temperate climates. Sci Rep. 2020;10(1):7990.

61. Rohousova I, Hostomska J, Vlkova M, Kobets T, Lipoldova M, Volf P. The protective effect against Leishmania infection conferred by sand fly bites is limited to short-term exposure. Int J Parasitol. 2011;41(5):481–5.

62. Kostalova T, Lestinova T, Sumova P, Vlkova M, Rohousova I, Berriatua E, et al. Canine Antibodies against Salivary Recombinant Proteins of Phlebotomus perniciosus: A Longitudinal Study in an Endemic Focus of Canine Leishmaniasis. PLoS Negl Trop Dis. 2015;9(6):e0003855.

63. Yared S, Gebresilassie A, Akililu E, Balkew M, Warburg A, Hailu A, et al. Habitat preference and seasonal dynamics of Phlebotomus orientalis in urban and semi-urban areas of kala-azar endemic district of Kafta Humera, northwest Ethiopia. Acta Trop. 2017;166:25–34.

64. Gidwani K, Picado A, Rijal S, Singh SP, Roy L, Volfova V, et al. Serological markers of sand fly exposure to evaluate insecticidal nets against visceral leishmaniasis in India and Nepal: a cluster-randomized trial. PLoS Negl Trop Dis. 2011;5(9):e1296.

65. Mahajan R, Owen SI, Kumar S, Pandey K, Kazmi S, Kumar V, et al. Prevalence and determinants of asymptomatic Leishmania infection in HIV-infected individuals living within visceral leishmaniasis endemic areas of Bihar, India. PLoS Negl Trop Dis. 2022;16(8):e0010718.

66. Quinnell RJ, Soremekun S, Bates PA, Rogers ME, Garcez LM, Courtenay O. Antibody response to sand fly saliva is a marker of transmission intensity but not disease progression in dogs naturally infected with Leishmania infantum. Parasit Vectors. 2018;11(1):7.

67. Ortuno M, Munoz C, Spitzova T, Sumova P, Iborra MA, Perez-Cutillas P, et al. Exposure to Phlebotomus perniciosus sandfly vectors is positively associated with Toscana virus and Leishmania infantum infection in human blood donors in Murcia Region, southeast Spain. Transbound Emerg Dis. 2022;69(5):e1854–e64.

68. Albuquerque A, Campino L, Cardoso L, Cortes S. Evaluation of four molecular methods to detect Leishmania infection in dogs. Parasit Vectors. 2017;10(1):57.

69. Galluzzi L, Ceccarelli M, Diotallevi A, Menotta M, Magnani M. Real-time PCR applications for diagnosis of leishmaniasis. Parasit Vectors. 2018;11(1):273.

70. Abbasi I, Aramin S, Hailu A, Shiferaw W, Kassahun A, Belay S, et al. Evaluation of PCR procedures for detecting and quantifying Leishmania donovani DNA in large numbers of dried human blood samples from a visceral leishmaniasis focus in Northern Ethiopia. BMC Infect Dis. 2013;13:153.

71. Chapman LAC, Morgan ALK, Adams ER, Bern C, Medley GF, Hollingsworth TD. Age trends in asymptomatic and symptomatic Leishmania donovani infection in the Indian subcontinent: A review and analysis of data from diagnostic and epidemiological studies. PLoS Negl Trop Dis. 2018;12(12):e0006803.

72. Sudarshan M, Singh T, Singh AK, Chourasia A, Singh B, Wilson ME, et al. Quantitative PCR in epidemiology for early detection of visceral leishmaniasis cases in India. PLoS Negl Trop Dis. 2014;8(12):e3366.

73. Bhattarai NR, Van der Auwera G, Khanal B, De Doncker S, Rijal S, Das ML, et al. PCR and direct agglutination as Leishmania infection markers among healthy Nepalese subjects living in areas endemic for Kala-Azar. Trop Med Int Health. 2009;14(4):404–11.

74. De Pascali AM, Todeschini R, Baiocchi S, Ortalli M, Attard L, Ibarra-Meneses AV, et al. Test combination to detect latent Leishmania infection: A prevalence study in a newly endemic area for L. infantum, northeastern Italy. PLoS Negl Trop Dis. 2022;16(8):e0010676.

75. Leveque MF, Lachaud L, Simon L, Battery E, Marty P, Pomares C. Place of Serology in the Diagnosis of Zoonotic Leishmaniases With a Focus on Visceral Leishmaniasis Due to Leishmania infantum. Front Cell Infect Microbiol. 2020;10:67.

76. Roberts T, Keddie SH, Rattanavong S, Gomez SR, Bradley J, Keogh RH, et al. Accuracy of the direct agglutination test for diagnosis of visceral leishmaniasis: a systematic review and meta-analysis. BMC Infect Dis. 2023;23(1):782.

77. Dey R, Alshaweesh J, Singh KP, Lypaczewski P, Karmakar S, Klenow L, et al. Production of leishmanin skin test antigen from Leishmania donovani for future reintroduction in the field. Nat Commun. 2023;14(1):7028.

78. Singh OP, Sundar S. Whole blood assay and visceral leishmaniasis: Challenges and promises. Immunobiology. 2014;219(4):323–8.

