## supplemental data for "Demographic characteristics and prevalence of asymptomatic *Leishmania donovani* infection in migrant workers working in an endemic area in Northwest Ethiopia"

**Table S1**

**Age of farmer and student FC and RC**

| | **Farmer**<br>Age (years) | **Student**<br>Age (years) |
|---|---|---|
| **FC** | 24 [20-32.3] | 19 [18-20] |
| **RC** | 26 [22-34] | 20 [19-23] |
| ***p* value*** | <0.0001 | <0.0001 |

Statistical difference between the ages of farmer FC (n=196) and RC (n=91) and student FC (n=304) and RC (n=77) was determined using a Mann-Whitney test. Data are presented as medians followed by IQR in square brackets. FC=first comer, RC=repeat comer.

**Table S2**

**Permanent place of residence**

| **Districts** | **FC** | **RC** |
|---|---|---|
| **Aderemet** | **0** | **1** |
| Aderkay | 3 | 2 |
| Amara Sayint | 0 | 1 |
| Andabet | 43 | 0 |
| Bahir Dar | 0 | 1 |
| **Belesa** | **10** | **5** |
| Beyeda | 8 | 0 |
| Burie | 0 | 1 |
| Dabat | 14 | 3 |
| Debark | 31 | 20 |
| Debre Markos | 0 | 1 |
| Dembia | 32 | 9 |
| Dera | 1 | 0 |
| **Ebenat** | **4** | **1** |
| Estie | 38 | 21 |
| Farta | 13 | 13 |
| **Fogera** | **3** | 4 |
| Gindewoin | 3 | 3 |
| **Gondar** | **8** | **4** |
| Jabitena | 2 | 4 |
| Janamora | 10 | 7 |
| Kuarit | 2 | 0 |
| **Libokemkem** | **16** | **9** |
| Makesegnit | 79 | 22 |
| Merawi | 27 | 10 |
| **Metema** | **0** | **1** |
| Negade Bahir | 0 | 1 |
| **Sanja** | **8** | **12** |
| Sekela | 0 | 1 |
| **Sekota** | **3** | **2** |
| Shawira | 15 | 0 |
| Shebel Berenta | 0 | 1 |
| Simada | 5 | 7 |
| Telemit | 133 | 8 |

The districts highlighted in bold indicate districts that are endemic for visceral leishmaniasis

**Table S3**

**Number of PCR cycles**

| **T1**<br>**FC # cycles** | | **T1**<br>**RC # cycles** |
|---|---|---|
| 37.8 | | 32.8 |
| 34.1 | | 36.8 |
| 36.9 | | 39.5 |
| 30.1 | | 38.9 |
| 38.5 | | 38.2 |
| 37.8 | | 37.1 |
| 37.7 | | 33.5 |
| 32.6 | | 27.3 |
| 35.1 | | 37.4 |
| 35.1 | | 34.6 |
| 37.8 | | 36.7 |
| 34.4 | | 37.8 |
| 35.3 | | 35.5 |
| 38.7 | | 36.2 |
| 37.5 | | 40.0 |
| **T2**<br>**FC # cycles** | | **T2**<br>**RC # cycles** |
| 34.6 | | 37.1 |
| 34.09 | | 38.75 |
| 39.25 | | 37.7 |
| 32.6 | | |
| 35.9 | | |
| 32.61 | | |
| 38.51 | | |
| 37.6 | | |
| 37.63 | | |

PCR was performed as described in Materials and Methods. The data represent the number of PCR cycles for each positive sample. FC=first comer, RC=repeat comer.

**Table S4**

| PCR | T1 | T2 |
|---|---|---|
| | 37.8 | Negative |
| | 37.8 | Negative |
| | 37.7 | Negative |
| | 37.8 | Negative |
| | 37.5 | Negative |

PCR was performed as described in Materials and Methods. The data represent the number of PCR cycles for each positive sample at T1 that tested negative at T2.

**Table S5.** Cohen's kappa values for pairwise agreement between different tests for all participants, and for first comers (FC) and repeat comers (RC), at timepoints 1 and 2 (T1 and T2)

| | **rK39 vs DAT** | | **rK39 vs PCR** | | **DAT vs PCR** | |
|---|---|---|---|---|---|---|
| **T1** | **n** | **Cohen's kappa** | **n** | **Cohen's kappa** | **n** | **Cohen's kappa** |
| All | 658 | 0.09 | 83 | -0.07 | 80 | 0.06 |
| FC | 495 | -0.01 | 47 | 0 | 46 | -0.04 |
| RC | 163 | 0.11 | 36 | 0.16 | 34 | 0.12 |
| **T2** | | | | | | |
| All | 154 | 0.23 | 42 | -0.18 | 42 | -0.22 |
| FC | 132 | 0.16 | 32 | -0.13 | 32 | -0.19 |
| RC | 22 | 0.56 | 10 | -0.25 | 10 | -0.25 |

**Table S6**. Fleiss' kappa values for three-way agreement between different tests for all participants, and for first comers (FC) and repeat comers (RC), at the first and second timepoints (T1 and T2)

| Group | n | Fleiss' kappa |
|---|---|---|
| **T1** | | |
| All | 80 | -0.03 |
| FC | 46 | -0.13 |
| RC | 34 | -0.01 |
| **T2** | | |
| All | 42 | 0.03 |
| FC | 32 | -0.03 |
| RC | 10 | 0.17 |

**Table S7**

**MW who developed VL by T2**

| | T1 rK39 | T2 rK39 | T1 DAT | T2 DAT | T1 PCR | T2 PCR |
|---|---|---|---|---|---|---|
| FC | positive | positive | <1/1600 | 1/25600 | nt | nt |
| RC | positive | positive | <1/1600 | 1/12800 | negative | 38.7 |

The trajectory of the different tests (rK39, DAT and PCR) between T1 and T2 for the two migrant workers who developed VL. FC=first comer, RC=repeat comer. nt=not tested

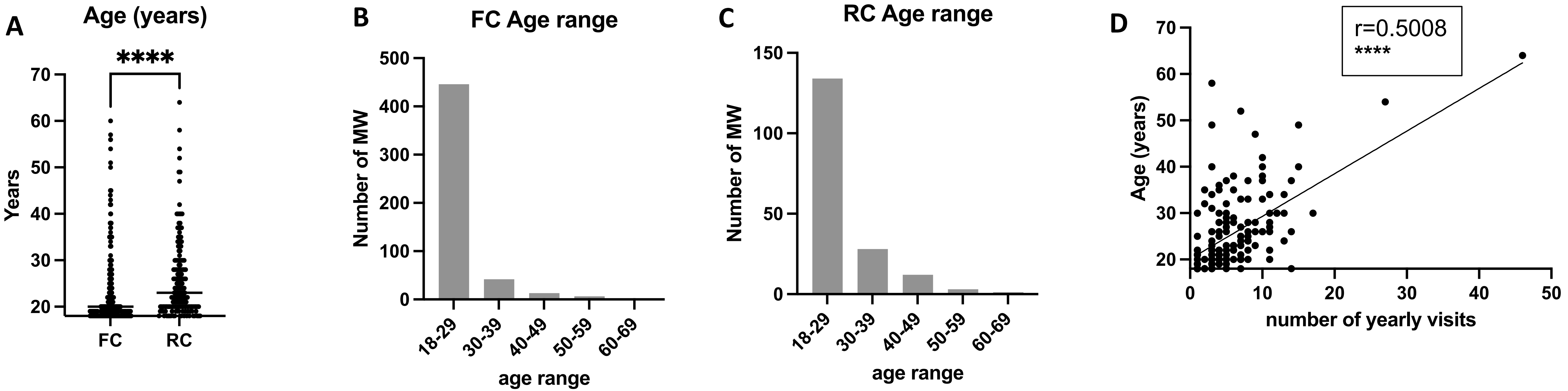


**Figure S1: Ages of the two groups of MW**

(A) Statistical difference between the ages of the FC (n=508) and the RC (n=175) was determined using a Mann-Whitney test. The straight line represents the median. (B) Number of FC and RC (C) per age range. (D) The correlation between the different ages of the RC and the number of times (in years) they visited the area of Metema/Abdurafi was measured using a Spearman test. FC=first comer, RC=repeat comer.

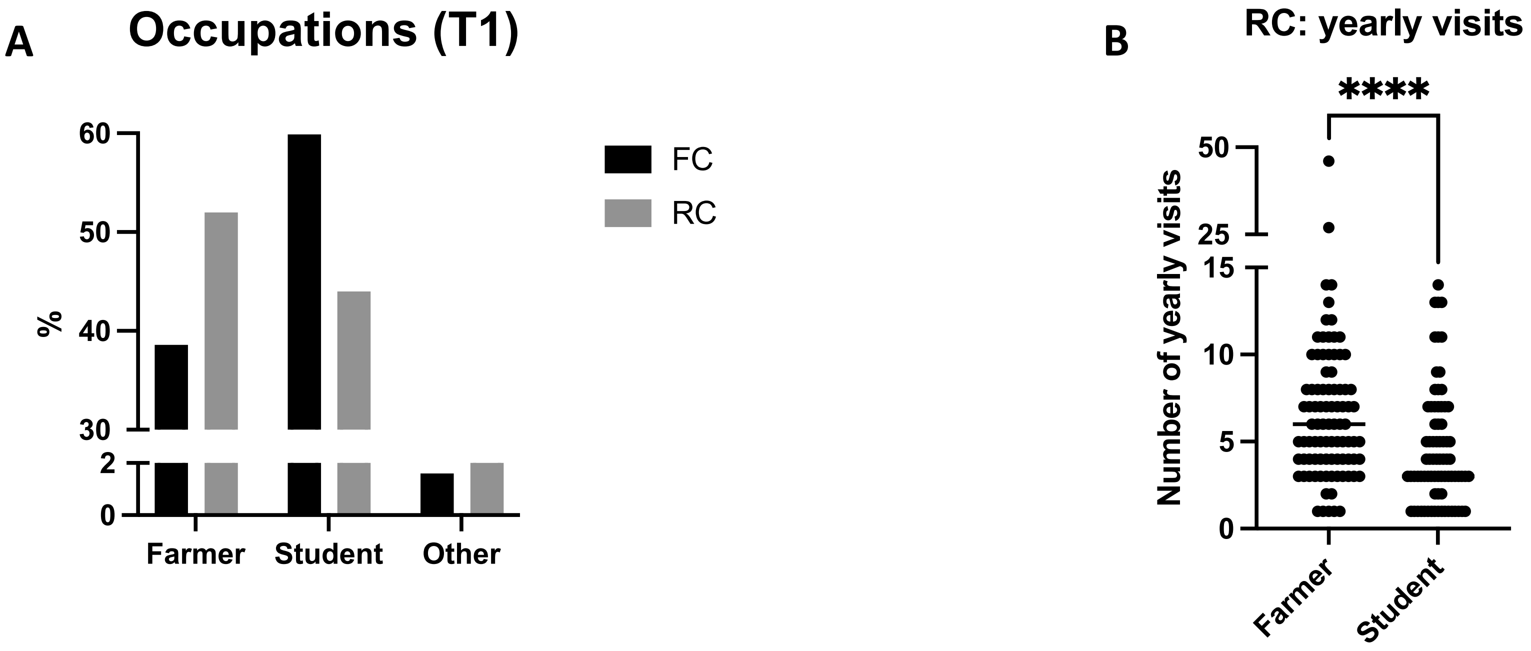


**Figure S2: Migrant workers' occupations**

(A) Percentage of FC and RC whose main occupation is farmer, student or other (defined as other than farmer or student). (B) Statistical difference between the number of times (in years) farmer (n=90) and student (n=77) RC visited the area of Metema/Abdurafi was measured using a Mann-Whitney test. The straight line represents the median. FC=first comer, RC=repeat comer.

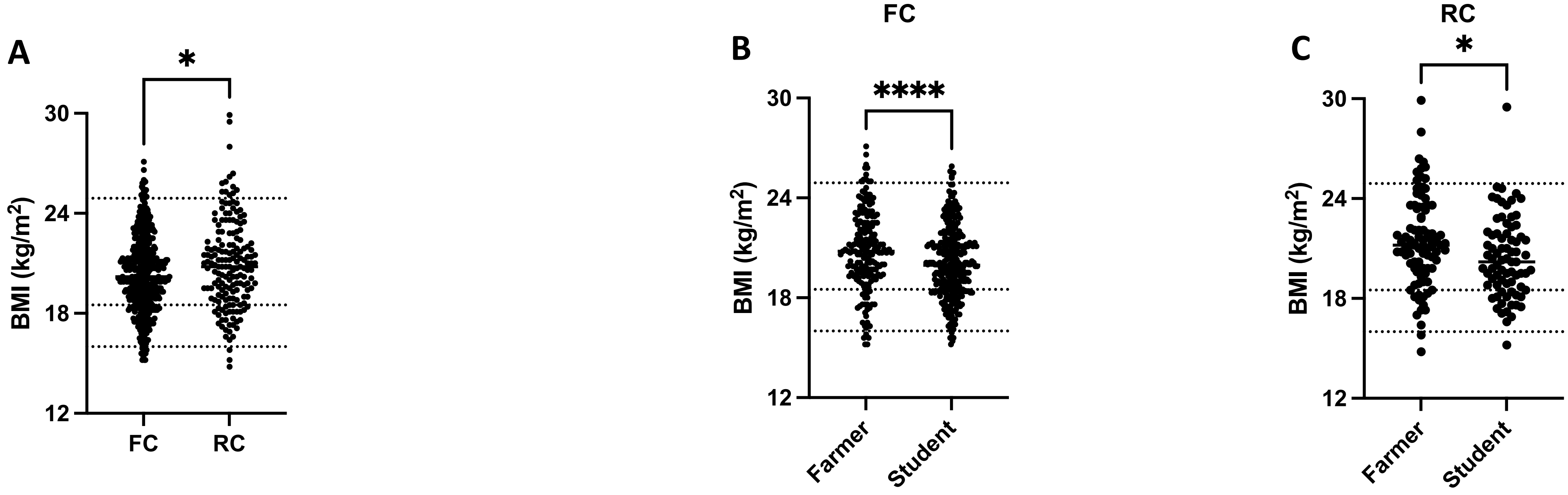


**Figure S3: Migrant workers' body mass index**

(A) Statistical difference between the body mass index (BMI) of FC (n=511) and RC (n=174) at T1 was measured using a Mann-Whitney test. Statistical differences between the BMI of FC who were farmers (n=197) or students (n=306)(B) or RC who were farmers (n=91) or students (n=77)(C) at T1 were measured using a Mann-Whitney test. The straight line represents the median. The lower dotted line is set at 16, the middle dotted line is set at 18.5 and the upper dotted line is set at 24.9. FC=first comer, RC=repeat comer..

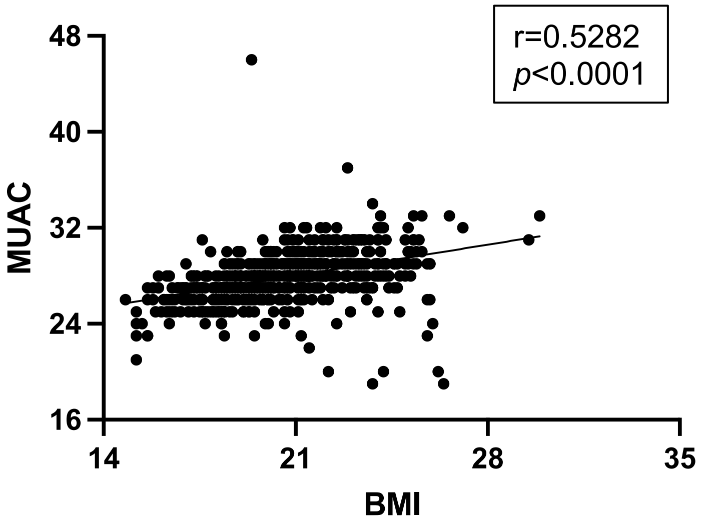


**Figure S4: Correlation between body mass index and middle upper arm circumference amongst MW**

The correlation between the BMI and MUAC at T1 was measured by a Spearman test (n=672).

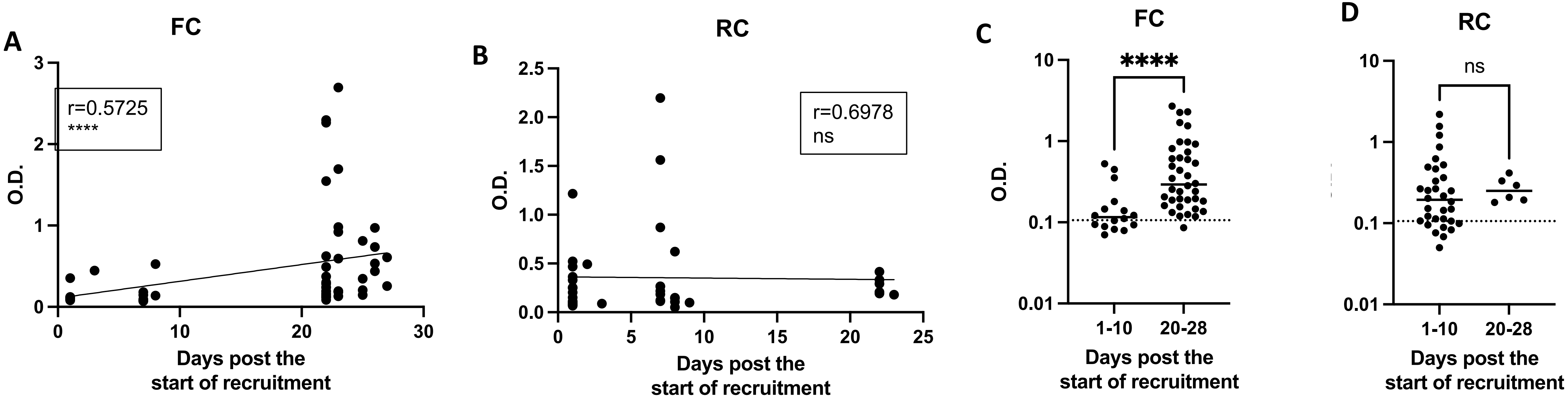


**Figure S5: Exposure to sand fly saliva**

An ELISA was performed to measure the levels of anti-*P. Orientalis* saliva antibodies in plasma as described in [44], the results are presented as optical density (O.D.). (A) (B) The correlations between the number of days post the start of recruitment and the O.D. for FC (A)(n=54) and RC (B)(n=38) was measured by a Spearman test. Comparison of FC (n=54), RC (n=38) and Healthy Non-endemic controls (HNEC, n=24). (C) Statistical difference between the O.D. of FC who were recruited during the first 10 days (n=16) or the last 8 days (n=38) at T1 was determined using a Mann-Whitney test. (D) Statistical difference between the O.D. of RC who were recruited during the first 10 days (n=32) or the last 8 days (n=6) at T1 was determined using a Mann-Whitney test.

The dotted line represents the cut-off value and the straight line represents the median. ns=not significant. FC=first comer, RC=repeat comer.

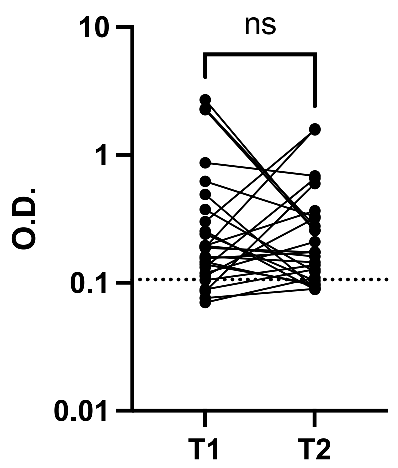


**Figure S6: levels of anti-saliva antibodies: longitudinal follow-up of MW**
An ELISA was performed to measure the levels of anti-*P. Orientalis* saliva antibodies in plasma as described in [44], the results are presented as optical density (O.D.). MW were followed longitudinally between T1 and T2 (n=26) and the statistical difference between these two time points was measured by a Wilcoxon matched-pairs test. The dotted line represents the cut-off value.

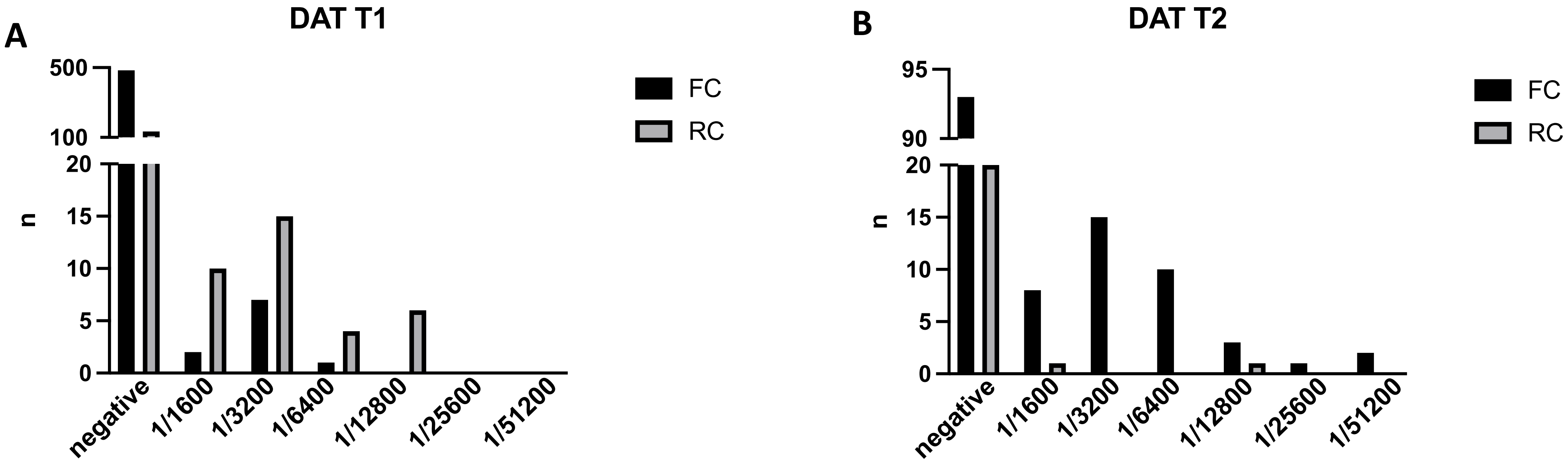


**Figure S7: DAT: number of negative tests and positive tests for each titre at T1 and T2**

Direct Agglutination Test was performed on dried blood spots as described in Materials and Methods. DAT results were grouped as negative (<1/1600) and positive (≥1/1600).

(A) The number of negative and those positive for each titre at T1 for FC (solid bars, n=495) and RC (grey bars, n=169). (B) The number of negative and those positive for each titre at T2 for FC (solid bars, n=132) and RC (grey bars, n=22).

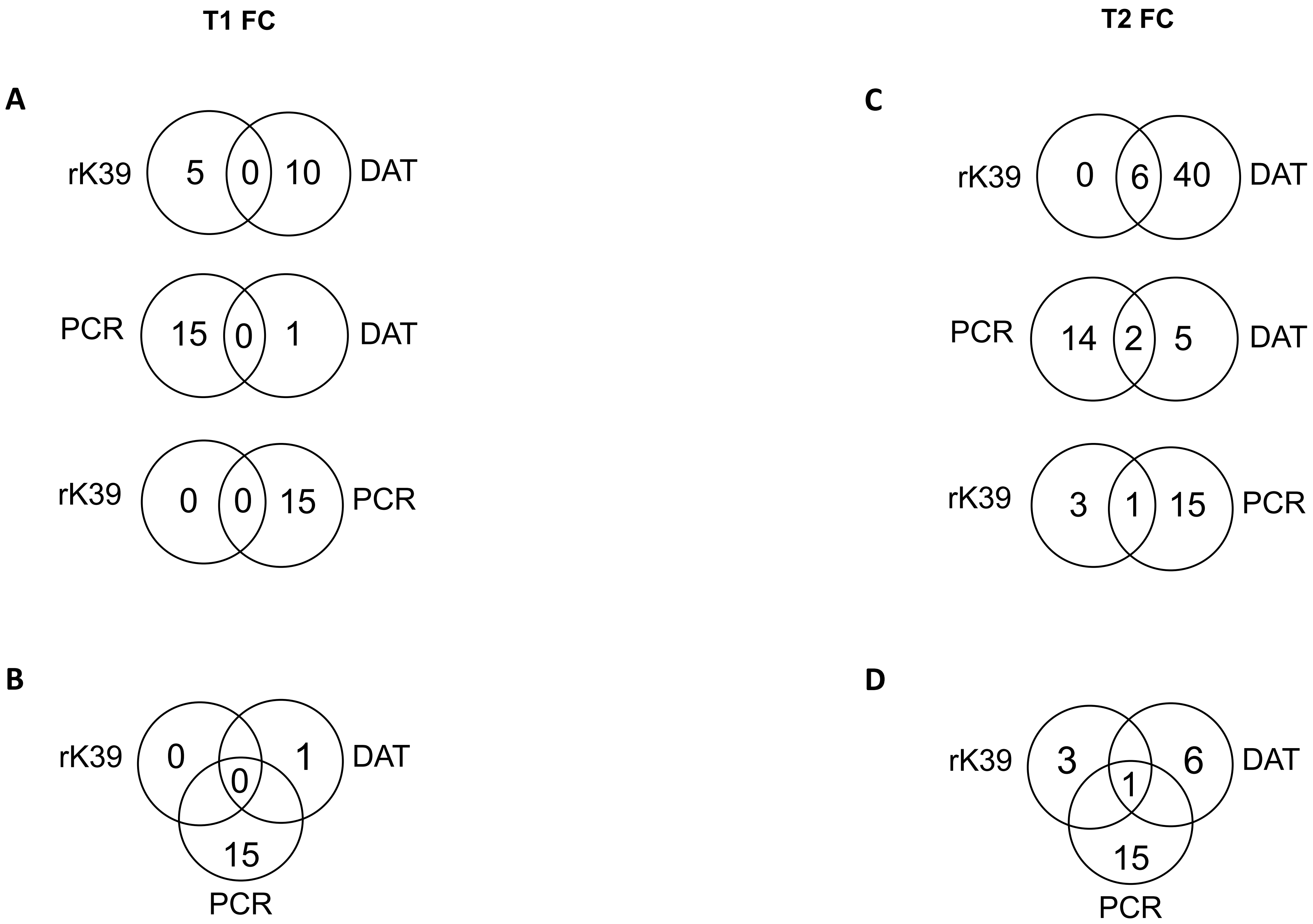


**Figure S8: Concordance between the different tests for FC**

Agreement between a combination of 2 tests at T1 (A) and T2 (C) or of three tests at T1 (B) and T2 (D).

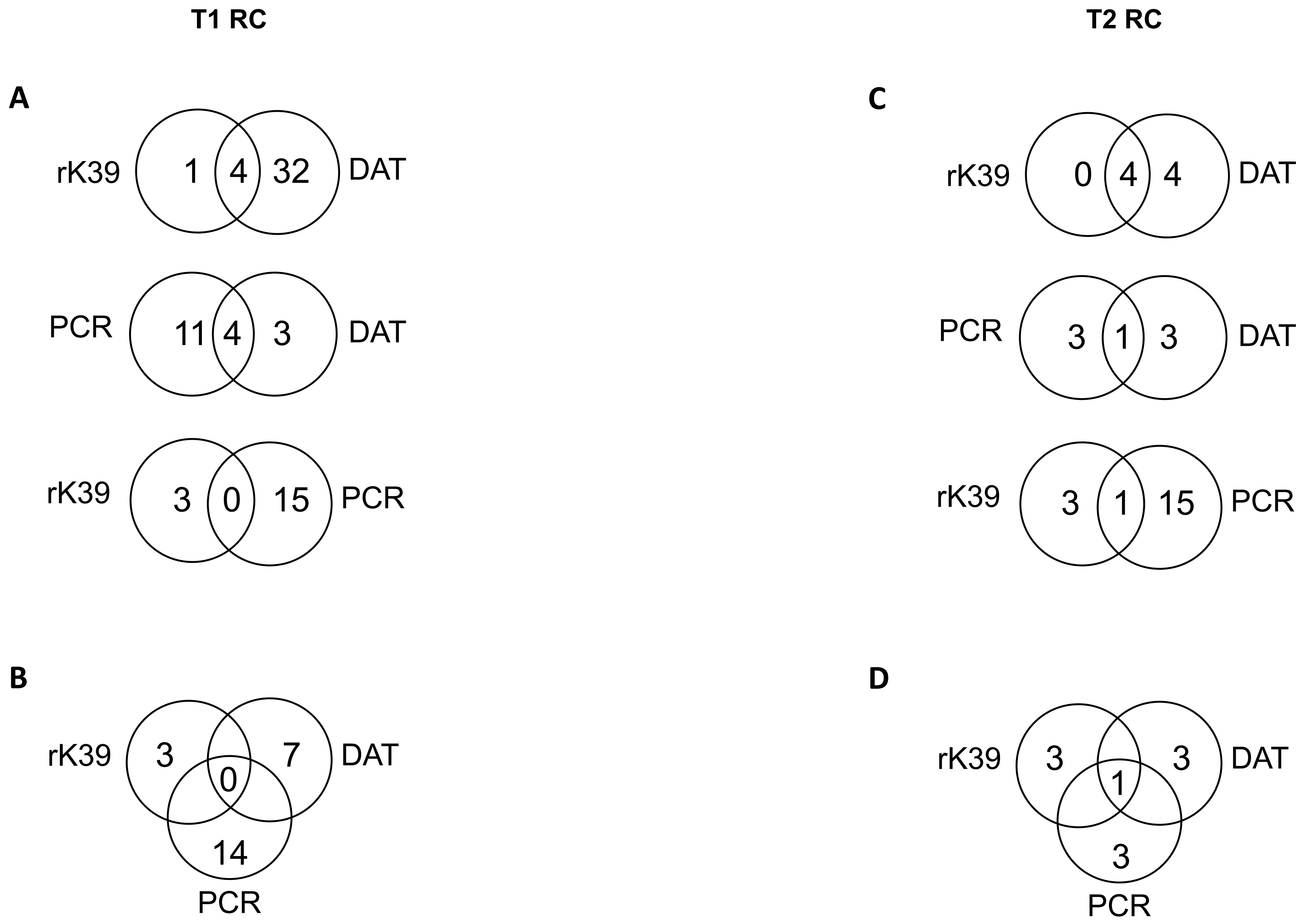


**Figure S9: Concordance between the different tests for RC**

Agreement between a combination of 2 tests at T1 (A) and T2 (C) or of three tests at T1 (B) and T2 (D).
